# A Comparative Analysis in a Clinical Cohort: Multiple Imputation by Chained Equations and a Novel Super Learner-Based Imputation Approach

**DOI:** 10.1101/2025.11.26.25341082

**Authors:** Tony Zbysinski, Lezhou Wu, Justin Dale, James Coates, Karan Sapiah, Jamie Reuben, Frank Markson, Ujjwal Kulkarni, Nazmul Islam

## Abstract

**Background:** Missing data is a challenge in clinical research, especially in real-world data (RWD), where complete case analysis can bias results and reduce power. Ensemble learning approaches like Super Learner (SL) show strong numerical performance for prediction problems, but their use for missing value imputation (MVI) in oncology datasets is unexplored. We sought to develop and evaluate a novel SL-based imputation function that can impute multiple variables and quantify observation-specific uncertainty.

**Methods:** We analyzed two independent cohorts of acute myeloid leukemia patients (n=1641), 546 patients from the University of Colorado and 1095 from an external real-world cohort. The SL-based MVI function includes data processing, predictor selection, binary and continuous variable pipelines, and automatic performance measurement. Ensembles for both binary and continuous variables integrate diverse base learners, such as generalized linear models, random forests, and neural networks, via a meta learner that optimizes predictive accuracy. The binary variable pipeline’s SL ensemble was optimized using area under the curve (AUC), while the continuous variable pipeline’s SL ensemble was optimized via non-negative least squares. Performance was compared to multiple imputation by chained equations (MICE) using balanced accuracy, F_1_-score, root mean square error (RMSE), and visualizations. Observation-specific uncertainty was quantified for all imputations of both binary and continuous variables, with both additionally having lower and upper resampling-based potential imputation values. The SL cross-validation loop, SL ensemble trained for imputation, and resampling all supported parallelization. Clinically significant features of the cohorts were selected *a priori* based on prior literature.

**Results:** In a numerical experiment with 9 clinically important binary features, the proposed MVI function imputed and achieved higher balanced accuracy than MICE for 7/9 variables (mean balanced accuracy 89.04% vs 80.75%) with comparable performance for the other 2 variables. The continuous variable SL ensemble, across 4 variables, showed an average 24.45% lower RMSE than MICE. On average, the SL ensemble trained for prediction took 145.02 seconds to process for binary targets.

**Conclusions:** This study demonstrates that the SL-based imputation function has improved performance over MICE in high-dimensional RWD while providing novel, observation-level uncertainty quantification.

## Background

Data missingness is a common occurrence in clinical research and poses a persistent threat to the validity of statistical inference and the strength of predictive models. Missing values can arise for several reasons such as instrument failure, patient dropout, or recording decisions driven by underlying patient characteristics (1,2). These challenges lead to reduced statistical power, biased effect estimates, and compromised precision, ultimately undermining predictive modeling efforts (3,4). As real-world data (RWD) sources grow in volume and complexity, particularly in high-dimensional settings such as electronic health records or genomic repositories, the challenge of appropriately handling missingness increases. Traditional approaches like complete-case analysis become unrealistic in many scenarios and can propagate selection bias (5).

Early work on missing data classified the mechanisms by which values are missing. These include missing completely at random (MCAR), missing at random (MAR), and missing not at random (MNAR). Such mechanisms are one part of understanding which imputation approach is needed (3). Classical single and multiple imputation techniques, such as mean/median/mode substitution or multiple imputation via chained equations (MICE), provide straightforward and satisfactory solutions under MCAR and MAR assumptions, but can perform poorly when these assumptions are violated or when variable interdependencies are complex (6,7). Machine learning (ML) methods including k-nearest neighbors (k-NN), random forests, support vector machines, and others have typically demonstrated improved performance over purely statistical methods, particularly for moderate missingness and MAR settings (7–9). Deep learning approaches, such as generative adversarial imputation networks and variational autoencoders, offer promise in capturing nonlinear relationships and high-order interactions (5,10), but comparative studies against well-established algorithms like missForest and MICE in RWD scenarios have shown that conventional methods remain competitive or superior in practice at this time. This is likely due to the volume of tuning parameters and the dataset size requirements of deep learning approaches (11).

Despite the broad array of available imputation strategies, no single method uniformly or consistently outperforms others across mechanisms, domains, and dimensionalities (12). In clinical settings, where variable distributions may be skewed and the mechanisms of missingness are frequently a complex mix of MCAR, MAR, and MNAR, ensemble learning techniques hold particular promise by combining multiple base learners to hedge against model-specific weaknesses (12). The Super Learner (SL) framework implements a stacking ensemble that uses cross-validation (CV) specific risk estimates, risk being a term used for brevity when referring to objective functions to optimally weight ML algorithms (e.g. minimizing mean squared error). In this approach it combines the strengths of multiple semi-parametric or non-parametric models to make a balanced single weighted imputation of missing values (13,14), unlike MICE which is a more iterative process. It has been applied and tested for missing value imputation (MVI) in previous literature, showing superior performance in at least synthetic data compared to more popular methods such as MICE (15). Outside of MVI, but within the clinical domain, SL-based approaches have yielded strong results for numerous prediction-related tasks (16–20). However, SL’s adoption for imputation in clinical RWD has been limited despite its ability to outperform traditional methods. This advancement is critical given the potential impact of imputation on downstream tasks, such as treatment effect estimation, risk stratification, and clinical decision-making.

This study addresses a key gap in application by evaluating the performance of MVI techniques using a SL-based approach on a high-dimensional dataset. In addition, we build on this approach by introducing a novel function designed to greatly simplify the imputation process with SL and provide measures of uncertainty for other modeling tasks. The evaluation is conducted on clinical data comprising acute myeloid leukemia (AML) patients treated with venetoclax plus azacitidine (Ven/Aza) or cytarabine plus anthracycline (7+3). We experiment with varying missingness ratios and mixed or unknown missingness mechanisms to reflect realistic challenges in clinical data analysis. By benchmarking our SL-based imputation function against a conventional statistical technique and selected ML methods, we aim to determine whether a SL-based approach can deliver more robust and accurate imputations under clinical RWD conditions and provide uncertainty measures for downstream tasks. Our specific objectives are threefold: 1. Develop a novel user-friendly SL-based imputation function capable of iteratively imputing multiple target variables while quantifying uncertainty measures for other modeling purposes; 2. Apply the SL-based imputation function to impute missing diagnostic patient AML data and quantify numerical performance of singular learners of the ensemble; 3. Compare the SL-based imputation function’s predictive performance to state-of-the-art MICE performance. Note the proposed model is disease-agnostic, much like most imputation approaches such as MICE, and thus can be applied beyond AML without loss of generalizability. While we confine our objectives to imputing baseline features for convenience, the function can be generalized and extended to imputing values at other clinically relevant time-points.

## Methods

### Data Description

This study leveraged data from two cohorts (n=1641) of adult patients (aged ≥ 18 years) with newly diagnosed AML. Briefly, these patients were treated with Ven/Aza or 7+3. The University of Colorado (CU) cohort included 546 patients that began treatment in a clinical trial between January 2015 and March 2024 in the UCHealth system. Data is continuously updated over time as follow-up progresses, where follow-up ends at date of death, loss to follow-up, or until the most recent follow-up update. We censor patients only to their last known alive date. The Real World Cohort (RWC) was derived from the nationwide Flatiron Health electronic health record-derived, de-identified, longitudinal database consisting of patient-level structured and unstructured data curated via technology enabled abstraction and was comprised of 1095 patients from 87 unique sites of care (21,22). Flatiron Health dataset patients had a minimum of two documented clinical visits between January 2014 and December 2023 and received the Ven/Aza treatment as frontline therapy. The de-identified data is sourced from approximately 280 cancer clinics across the United States, including 800 sites of care primarily within community oncology settings. Inclusion and exclusion criteria were applied to minimize systematic differences between the cohorts, as discussed by Islam et al (23). Specifically, patients were excluded if they had acute promyelocytic leukemia, biphenotypic leukemia, chronic myeloid leukemia in blast crisis, or central nervous system involvement at diagnosis. A more thorough description of this data can be found in previous literature (23). Additional file 1, Figure S1, describes via a visual summary the genetic alteration patterns of both the RWC and the CU cohorts. Figure 1 illustrates the 5 key steps of our novel SL-based imputation framework, providing an overview of the processes detailed in the following sections on specification and data processing, imputation pipelines, performance assessment, and our application in high-dimensional clinical data.

**Figure 1.**
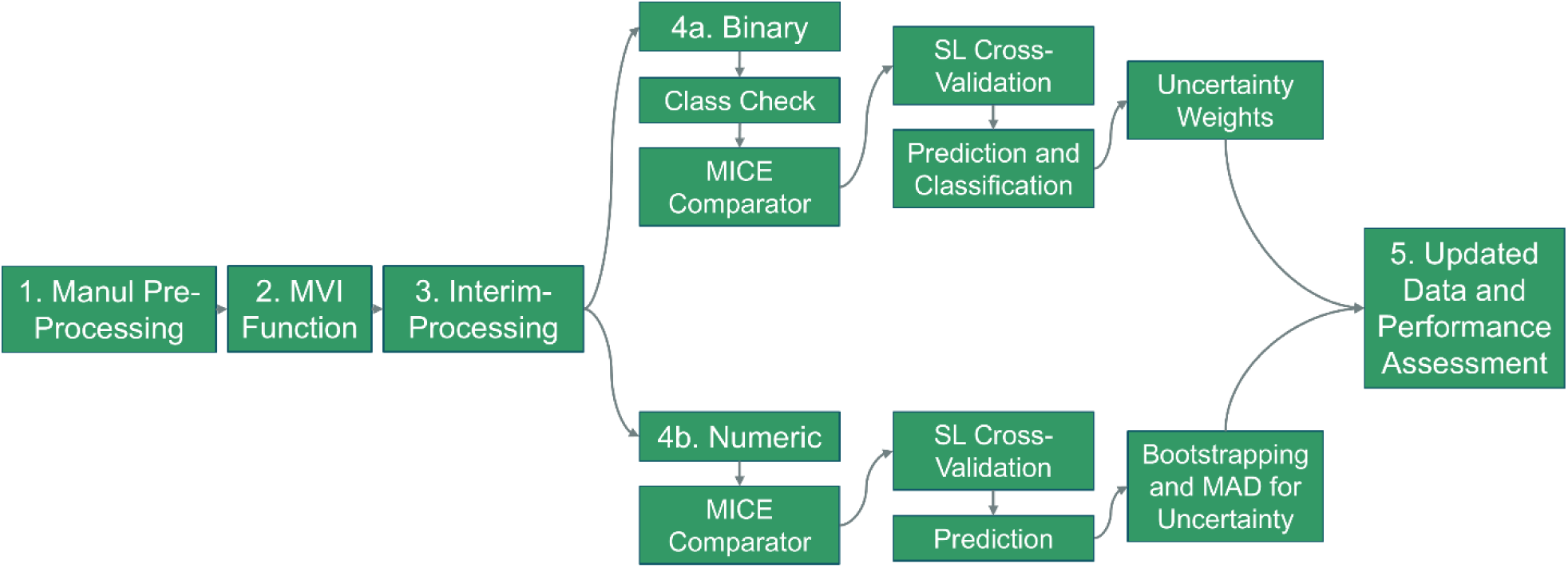
SL-based MVI imputation function methods flow diagram. *Abbreviations:* Median Absolute Deviation (MAD), Multiple Imputation by Chained Equations (MICE), Missing Value Imputation (MVI), Super Learner (SL).

### Specification and Data Processing

Step 1 includes all manual data pre-processing for the purpose of having correct data format and structure for the developed SL-based imputation function. Data pre-processing included, for our analysis, only variables measured at baseline (e.g., cytogenetic testing or laboratory measured values at diagnosis or treatment start date). The function accepts input data containing continuous and binary variable types; multinomial and ordinal features need to be transformed into a design matrix with binary columns before applying the MVI function. In our application, diagnostic clinical variables were harmonized between cohorts by merging on identified common variables. All dichotomous variables were made binary following traditional conventions for reference groups, coding values such as “No” or “Wild Type” as 0. To enhance imputation performance and reduce noise, variables considered irrelevant to modeling, including patient identifiers, upper and lower reference ranges for laboratory values, unknown data flags, and non-binary or non-continuous variables, were excluded during preprocessing, leaving 124 variables.

Step 2 specifies the SL-based imputation function and its interim preprocessing steps. This function accepts 4 parameters. The target_vars parameter takes a vector of binary or continuous variables for imputation. We recommend ordering these variables from least to most missingness, or from least to most importance, as the function’s iterative imputation scheme can leverage earlier imputations to improve subsequent estimates. The learners and screeners parameters are user-defined character vectors listing the base learners and screening wrappers to be incorporated into the SL calls. A comprehensive list of available wrappers can be retrieved using the listWrappers function in the SuperLearner package (13). The final parameter, df_input, specifies the input dataset, formatted according to the structure described in step 1.

In step 3, interim processing, the SL-based imputation function constructs the SL library, removes variables produced by earlier imputations that are irrelevant for subsequent targets, identifies the top predictors for each target variable, handles missingness in those predictors, and identifies the target variable as binary or continuous. An embedded utility function generates all possible combinations of user-specified base learners and screening wrappers. For example, if conditional forest and neural network learners are selected alongside correlation– and t-test-based screeners, each learner is paired with each screener to create a SL library of four unique algorithm inputs. It is important to note that the function has a built-in method of identifying and ranking top potential predictors based on missingness and correlation, reducing the demand for screener specification. To enhance interpretability in downstream outputs, a mapping of learner names is also established. If a user does not specify any base learning or screening wrappers, a default library of 19 learners is used with no screeners. These learners are listed with a brief description and reference in additional file 2, Table S1. While users may incorporate most available base learner wrappers and screeners within the SL framework, doing so may quickly become computationally infeasible, particularly in high-dimensional settings. Predictor selection is performed by first excluding variables generated during previous iterations (e.g., row ID trackers, uncertainty weights, confidence interval outputs) that do not inform estimation. For the remaining candidate predictors, a predictor score is computed as the product of (i) the absolute Pearson correlation with the target variable and (ii) the proportion of observed (non-missing) data, elevating predictors that are both informative and well-represented in the dataset. By default, the top 10 predictors ranked by this score are selected, keeping the next steps informed, parsimonious, and reducing overfitting. The number of predictors to use in modeling is a customizable parameter. Since the SL framework requires complete predictor data and missingness is prevalent in RWD, simple internal imputation is applied, median imputation for continuous variables and mode imputation for binary variables. This simple imputation step is used solely within the function and is not returned in the final output. Target variables with >80% missingness are flagged, and then the top predictor’s and target variable’s data is subset for subsequent SL-based methods to reduce dimensionality.

### SL-based Binary Imputation Pipeline

Step 4a defines the binary target variable pipeline, encompassing several key processes: (i) class balance checks, (ii) performance benchmarking via MICE, (iii) the core SL prediction process with CV, (iv) optimal threshold selection for classification, (v) resampling for accuracy variability, (vi) imputation of missing data, and (vii) generation of uncertainty weights. A class balance check identifies severely imbalanced binary variables. If one class comprises more than 92.5% of observations, or a class has equal to or less than 20 observations, a flag is triggered, and mode imputation is applied instead of SL-based modeling, as extreme imbalance can cause k-fold cross-validation to fail. In this case, a simple weight is assigned equal to the observed class proportion to reflect reduced uncertainty. For performance benchmarking of MICE, 30% of observed values are randomly masked, and MICE, using 5 imputations, is applied using a predictor matrix built from the input data. This provides a reference for comparison against the SL-based approach. The SL process then performs nested modeling and validation, where each base learner is trained and validated within cross-validation folds to generate predictions. These predictions are then combined by the meta-learner to optimize ensemble performance and minimize error (13). Within this pipeline, each learner undergoes 10-fold CV to estimate accuracy, and an outer CV loop (via CV.SuperLearner) evaluates the entire SL estimation process to provide performance estimates. The model uses the top 10 predictors identified in step 3, with simple internal imputation applied to handle missingness, and optimizes predictions for the area under the ROC curve (AUC). We next find the optimal threshold for classifying predicted probabilities by maximizing AUC when class imbalance is below 90%. In scenarios of 90-92.5% class imbalance, the F_1_-score was prioritized over AUC as it better reflects the trade-off between precision and recall and provides a more informative measure of performance for the minority class. Estimated probabilities obtained from the CV process are resampled 1000 times with replacement. This resampling enables calculation of confidence intervals for balanced accuracy, quantifying between subject variability in classification performance by applying the same threshold to each bootstrap replicate. Additionally, by resampling from the predicted distribution, computational feasibility is increased while accounting for uncertainty to an extent. The CV.SuperLearner object cannot directly estimate unseen data, we therefore rerun the SL process using snowSuperLearner with identical arguments to generate imputations for truly missing values.

### SL-based Binary Imputation Uncertainty

A similar 1000 resampling scheme is applied and the 2.5th and 97.5th percentiles of these samples form 95% observation-specific uncertainty intervals, reflecting variability in potential imputed predicted probabilities. The previously determined optimal threshold is applied to classify the original predicted probability and the lower 2.5th and upper 97.5th predicted probabilities. Finally, uncertainty weights are assigned to each imputed observation’s predicted probabilities to reflect classification confidence. These weights are calculated, as seen in Equation 1, by taking the absolute difference between the predicted probability of each observation, p^_i_ and the optimal threshold τ, scaled by the maximum possible distance to that threshold.

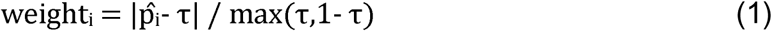

Predicted probabilities closer to the threshold receive lower weights, reflecting higher uncertainty, while those further away approach a weight of 1, normalizing uncertainty values between 0 and 1 while observed data is assigned a weight of 1.

### SL-based Continuous Imputation Pipeline

Step 4b describes the continuous target variable pipeline, which mirrors the binary pipeline but is adapted for continuous outcomes. After selecting the top 10 predictors and applying simple internal imputations to handle missingness among these predictors, a MICE-based comparison is conducted with specifications consistent with Step 4a. The SL CV process is then performed using 10-fold CV, a Gaussian family, and a non-negative least squares (NNLS) loss function to estimate performance for each base learner and the ensemble. The final SL ensemble model is subsequently trained on all observed data for the selected predictors and used to impute missing values in the target variable, producing direct value imputations rather than probabilities.

### SL-based Continuous Imputation Uncertainty

Imputation uncertainty for each imputed observation is quantified using a residual-based bootstrap approach, generating 1000 resampled imputations by adding resampled residuals to model-predicted values. To account for heteroskedasticity, the bootstrap resampling process selects the 50 residuals closest to each predicted value based on Euclidean distance from the observed target values, ensuring that resampled errors are drawn from observations with similar outcome magnitudes. The 2.5th and 97.5th percentiles of these samples form 95% observation-specific uncertainty intervals, reflecting variability in potential imputed values. Observation-specific uncertainty weights for continuous variables are shown in equation 2, calculated as the inverse of 1 plus the median absolute deviation (MAD) of the bootstrap sample distribution per observation, B_i_, normalized by the mean predicted value of the observation-specific bootstrap distribution pˉ_i_, thereby assigning higher weights to imputed values with lower relative variability.

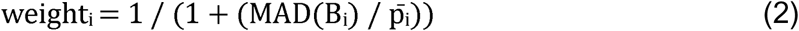

These outputs provide both a point estimate and observation-specific uncertainty intervals, enabling other analyses to account for uncertainty and down-weight less certain imputations.

### Performance Assessment and Application

Step 5 involves data output, iterative imputation, and comprehensive performance assessment. The SL-based imputation function produces an interim dataset in which each target variable is sequentially replaced by its imputed counterpart, supplemented with observation-specific uncertainty weights. The output additionally includes observation-specific 95% estimation uncertainty intervals that capture model uncertainty, enabling downstream analyses to consider a plausible range of imputed values rather than relying on single-point estimates. This augmented dataset is iteratively updated as each variable in the input vector is imputed, ultimately yielding a complete dataset supported with uncertainty-aware outputs. In addition to the final imputed data, the proposed imputation function returns a suite of diagnostic metrics and visualizations, providing strong transparency into the imputation process over standard methods like MICE. These include visualizations of top predictor scores for each target variable, flags for variables with >80% missingness, benchmark MICE performance metrics (confusion matrices and F_1_-scores for binary variables, RMSE, root mean square error, for continuous variables), and illustration of average CV risk estimates for every base learner in the SL ensemble (risk being RMSE for continuous variables, AUC for binary variable). The function also reports ensemble-level performance measures, balanced accuracy with bootstrap-based confidence intervals for binary targets, optimal thresholds for classification, and visualization of average ensemble weights based on nested CV folds illustrating each learner’s contribution to the SL model. Predicted values or probabilities are visualized along with resampling distributions. Furthermore, computing time is provided for both the independent MICE run and the final SL ensemble that is trained for imputation. Collectively, these outputs deliver not only a fully imputed dataset but also a novel, uncertainty-aware, and performance-benchmarked framework, offering users enhanced interpretability and awareness in the resulting imputations compared to conventional MVI approaches.

We summarize general descriptives of our merged 1641 patient cohort via counts for categorical variables and median and IQR for continuous variables. Additionally we describe missingness of the entire cohort leveraging the naniar R package (24). Missingness descriptives include missingness prevalence and visualizations to aid in identification of missingness patterns. In our initial testing application of this novel SL-based imputation function, we selected variables that have been previously identified in analyses of our data as high priority for upfront risk stratification and clinical relevance (23). These binary flag variables include NPM1 mutation, IDH1 mutation, IDH2 mutation, good risk cytogenetics as based on SWOG (25), inv(3) abnormality, TP53 mutation, –17 abnormality, t(9;11) abnormality, –5 abnormality, complex cytogenetics, and FLT3-ITD mutation. Given that these variables do not include a continuous data type, we additionally selected the laboratory-measured lactate dehydrogenase (LDH) variable for continuous variable evaluation due to its prognostic importance (26), with additional continuous variables of laboratory measured hemoglobin, platelets, and absolute neutrophil count (ANC). We ordered all selected variables from least to most missingness and supplied this vector to the MVI function, reporting summarizations of the previously mentioned performance metrics. In our application, we used the default 19 learner algorithms library. This diverse selection of learners ensures that the SL library balances simple benchmarks, linear and nonlinear models, tree-based approaches, penalized regression, and modern machine learning methods, enhancing flexibility and robustness in the estimation procedure and accounting for algorithmic uncertainty. To enable deeper insight into the SL-based imputation’s behavior and output data, we further isolated TP53 mutation and LDH to conduct a more detailed evaluation of SL predictive performance. This deeper comparison examined the performance of individual base learners, highlighting one algorithm that emerged as the top-performing single learner, referred to as the discrete learner. For SL processes, parallelization was configured using the parallel R package, and for resampling parallelization was configured using the future R package (27), both leveraging maximum cores. Lastly, to demonstrate how the SL-based imputations may compare to the MICE-based imputations in downstream work, we applied risk stratification logic, methodology detailed in a previous publication (28), to the cohort using these risk stratification variables. Differences between the imputation strategies’ influences on risk stratification were explored via the proportion of patients changing risk stratification, the change in median overall survival (OS) days for different risk stratifications, and Kaplan Meier survival curve visualizations. We also evaluated the prognostic utility of the derived risk stratifications using multivariable Cox proportional hazards models for overall survival, time from diagnosis to death or last follow-up, adjusting for age at diagnosis. Separate models were fit using the three-level risk stratification variable constructed from either MICE– or SL-imputed data, with the favorable category as the referent. All statistical analyses were performed in R version 4.4.1 (29). This study follows STROBE guidelines for reporting observational research (30).

## Results

### Descriptive Results

General cohort and target variable descriptive statistics are available in Table 1. Most of the 1641 patient cohort were male (703, 57.16%), White (1211, 73.8%), not Hispanic or Latino (1302, 79.34%), deceased at follow-up (1042, 63.5%), on the Ven/Aza treatment regime (1454, 88.60%), and are de novo AML (1076, 65.57%). Median age at enrollment was 74 (IQR 67-79). The target variables had missingness ratios that ranged from as low as 0.18% for good risk cytogenetics (3/1641) to high missingness of 54.17% for LDH (889/1641). Additional file 3, Figure S2 displays missingness ratios across all variables in the merged dataset, missingness across all variables can be summarized as highly diverse. Broadly, most variables were binary and had 40-80% missingness. Additional files 4-6, Figures S3-5, demonstrate missingness feature categories of cytogenetics, mutations, and lab values, respectively. Taken together, these missingness figures indicate primarily arbitrary and block patterns of missingness.

**Table 1.**
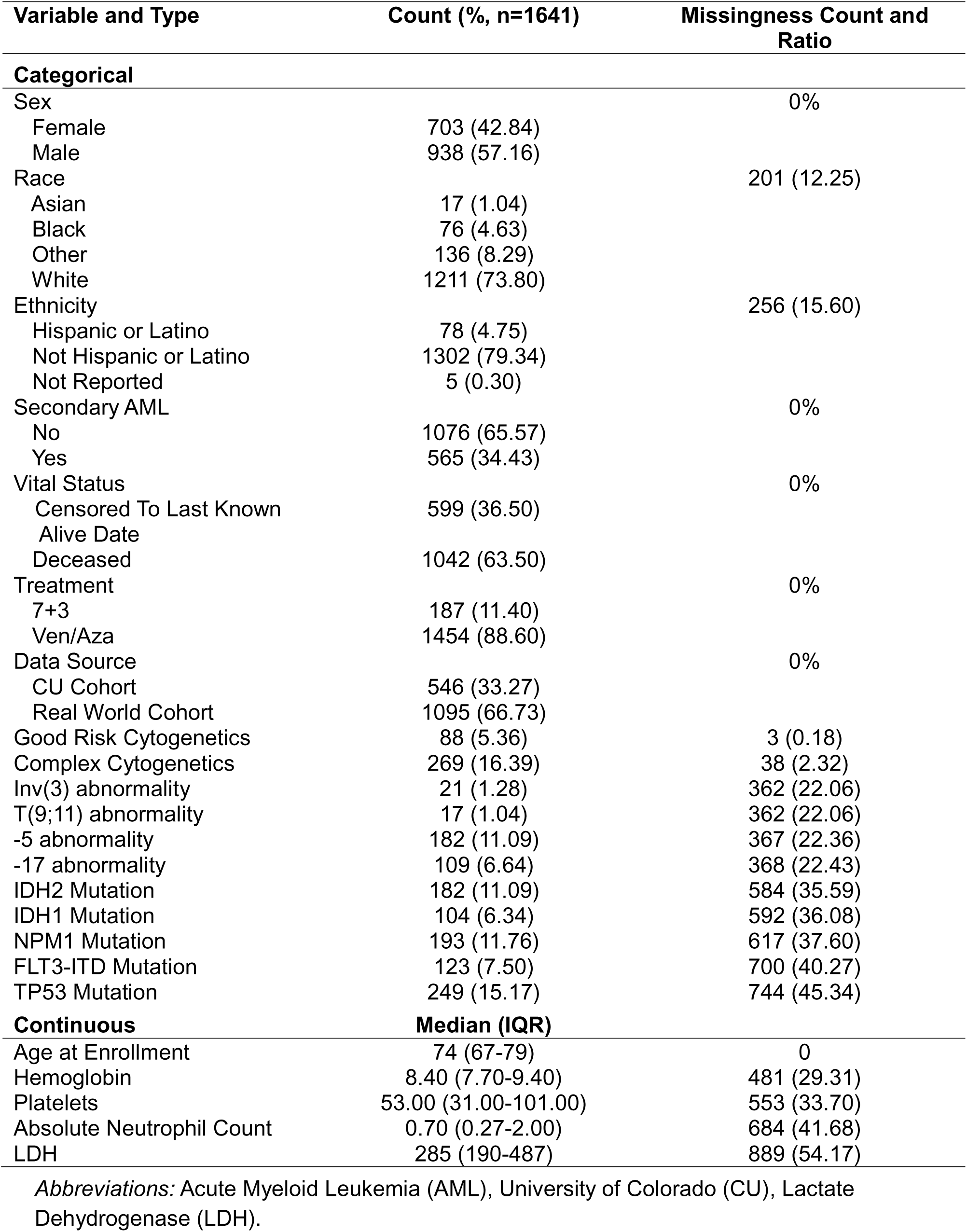
Merged cohort and target variable descriptive results and missingness.

### Primary Results

Table 2 details the performance for nearly all binary target variables of interest, comparing the performance of the SL-based ensemble approach to MICE. Variables inv(3) and t(9;11) abnormalities are not shown as they had high class imbalance and as a result were defaulted to the simple imputation pipeline. Performance measures included balanced accuracy, F_1_-score, and computational time. The SL-based ensemble approach had a greater mean balanced accuracy, 89.04%, than MICE, 80.75% and had superior balanced accuracy for 7 out of 9 variables, with 2 out of 9 variables having comparable (<1% difference) balanced accuracy. The mean F_1_-score for MICE, 94.34%, and the SL-based ensemble approach, 96.25%, were more mixed and slightly favoring the SL-based ensemble approach. Lastly, computational time was on average longer for the SL ensemble trained for imputation, 145 seconds, compared to MICE, 0.5 seconds.

**Table 2.** Performance of all binary target variables of interest compared between MICE and the SL-based imputation function approach.

| Variable | MICE<br>Balanced<br>Accuracy<br>(%) | SL<br>Balanced<br>Accuracy (95%<br>CI<sup>a</sup>, %) | MICE<br>F<sub>1</sub>-<br>score<br>(%) | SL F<sub>1</sub>-<br>score<br>(%) | MICE<br>Time<br>(seconds) | SL-based<br>Time<sup>b</sup><br>(seconds) |
| --- | --- | --- | --- | --- | --- | --- |
| Good Risk<br>Cytogenetics | 92.68 | 96.93 (94.11-99.15) | 98.91 | 99.61 | 0.79 | 137.67 |
| Complex<br>Cytogenetics | 95.04 | 98.88 (98.45-99.26) | 98.08 | 98.86 | 0.72 | 203.89 |
| -5 abnormality | 63.1 | 82.69 (79.89-85.36) | 90.47 | 87.82 | 0.42 | 190.88 |
| -17 abnormality | 58.03 | 57.58 (54.38-61.16) | 92.53 | 95.98 | 0.55 | 116.01 |
| IDH2 Mutation | 69.63 | 94.93 (92.87-96.63) | 88.93 | 97.52 | 0.51 | 131.08 |
| IDH1 Mutation | 95.99 | 95.19 (92.05-97.69) | 98.57 | 99.47 | 0.46 | 178.63 |
| NPM1 Mutation | 86.69 | 93.65 (91.54-95.71) | 95.95 | 97.22 | 0.48 | 123.61 |
| TP53 Mutation | 91.78 | 95.27 | 94.68 | 98.02 | 0.44 | 127.77 |
| FLT3-ITD<br>Mutation | 87.22 | 95.67 (93.12-97.81) | 90.91 | 91.71 | 0.53 | 95.65 |

<sup>a</sup> 95% confidence intervals represent the SL CV loop's 1000 resamples with classified via the optimal threshold for measures of precision for balanced accuracy.
<sup>b</sup> SL-based time refers to only the SL model trained for prediction and not the CV loop for performance measures.
*Abbreviations:* Multiple Imputation by Chained Equations (MICE), Super Learner (SL), Cross-Validation (CV).

In our specific performance analysis of the SL-based ensemble and MICE approaches of TP53 mutation and LDH imputation, we found that the SL-based approach was mostly superior to MICE. For TP53 mutation imputation, MICE had a balanced accuracy of 91.8%, sensitivity 93.7%, specificity 89.9%, and an F_1_-score of 94.7% while the SL-based ensemble approach had respective values of 95.3%, 99.4%, 91.2%, and 98.0%. Compared to the SL-based ensemble approach, the discrete learner of the ensemble had comparable performance (balanced accuracy 95.3%; sensitivity 99.4%; specificity 91.2%; F_1_-score 97.9%) and another select algorithm for comparison, GLM interaction, had worse performance (balanced accuracy 91.7%; sensitivity 96.6%; specificity 87.8%; F_1_-score 95.8%). Additional file 7, Figure S6, shows the top 10 predictors of the TP53 mutation variable that were used in the SL-based approach, indicating that several previous target variables that were imputed supported the imputation of TP53 mutation. Figure 2 shows the CV-specific base learner risks, in other words individual model performance, for TP53 mutation, suggesting that most base learners performed well (>0.9 AUC) with conditional forest having the absolute minimum risk observed. Figure 3 demonstrates the subsequent average assigned weights of the SL-based ensemble’s learners where many of the learners can be perceived to receive comparable weights as the SL technique aims to reduce overfitting. Lastly, for TP53 mutation, Figure 4 shows the predicted probability and corresponding resampled distribution of predicted probabilities based on 1000 resamples. With a calculated optimal threshold of 0.38 and the observed predicted probabilities, we would estimate that most classifications were 0 and were likely assigned moderate uncertainty weights. This is validated when analyzing the distribution of the TP53 mutation uncertainty weights (median 0.52, IQR 0.45-0.53). With respect to LDH, results were more mixed. MICE had a RMSE of 597, higher than that of the SL-based ensemble’s RMSE of 582. Furthermore, the discrete learner of the ensemble performed best with 557 RMSE, and other algorithms including neural network and all GLM algorithms, performed far worse with >5000 RMSE. Additional file 8, Figure S7, displays the top 10 predictors of the LDH variable that were used in the SL-based approach. Figure 5 shows the average CV-specific base learner risks for LDH. The SL-based ensemble’s learner’s average weights are shown in Figure 6, supporting a narrower range of base learners unequally.

**Figure 2.**
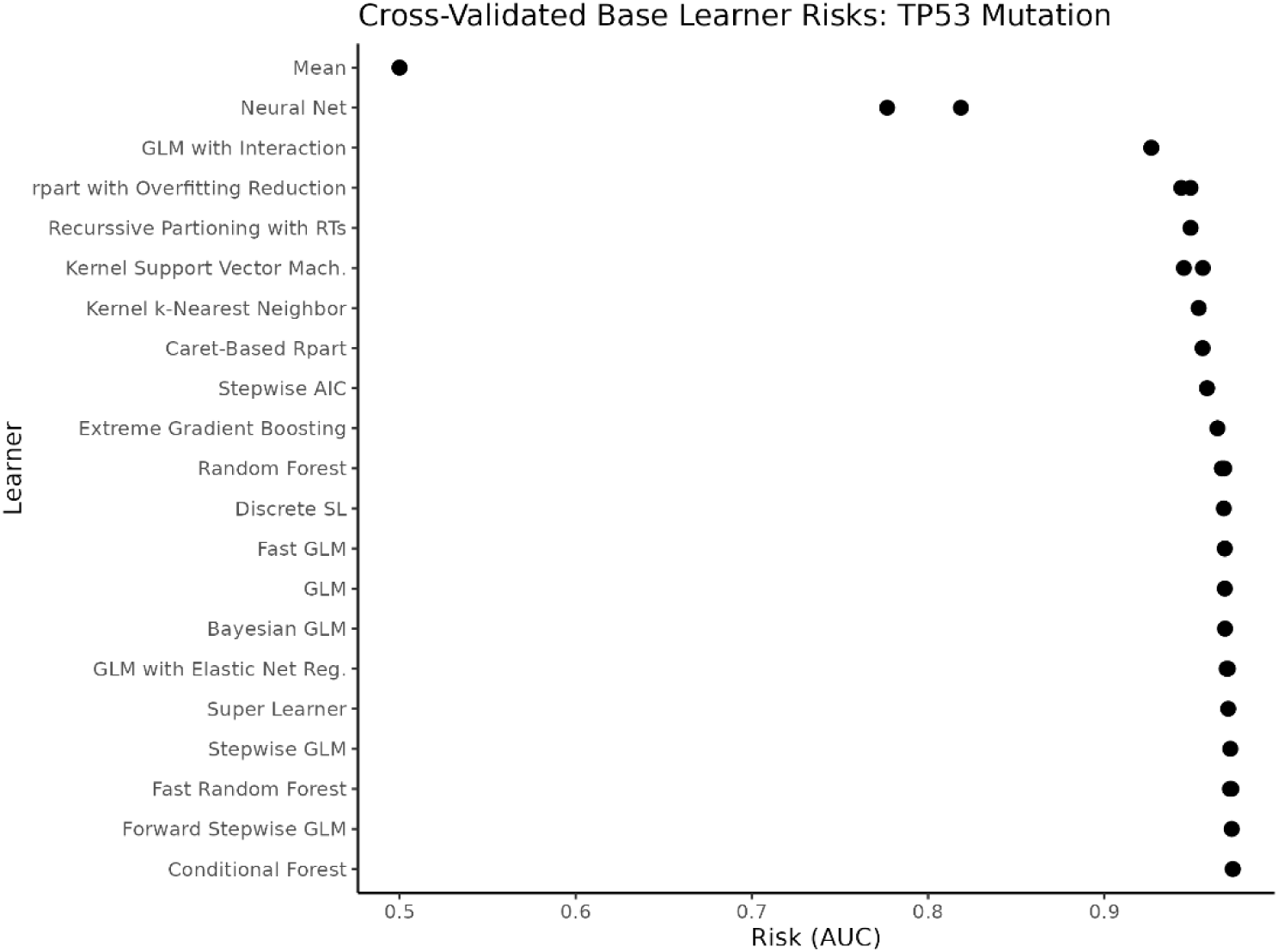
Cross-validated base learner risks for binary TP53 mutation variable prediction, risk measured as AUC. *Note:* multiple points may appear because of multiple default hyperparameter settings. *Abbreviations:* Area Under the Curve (AUC).

**Figure 3.**
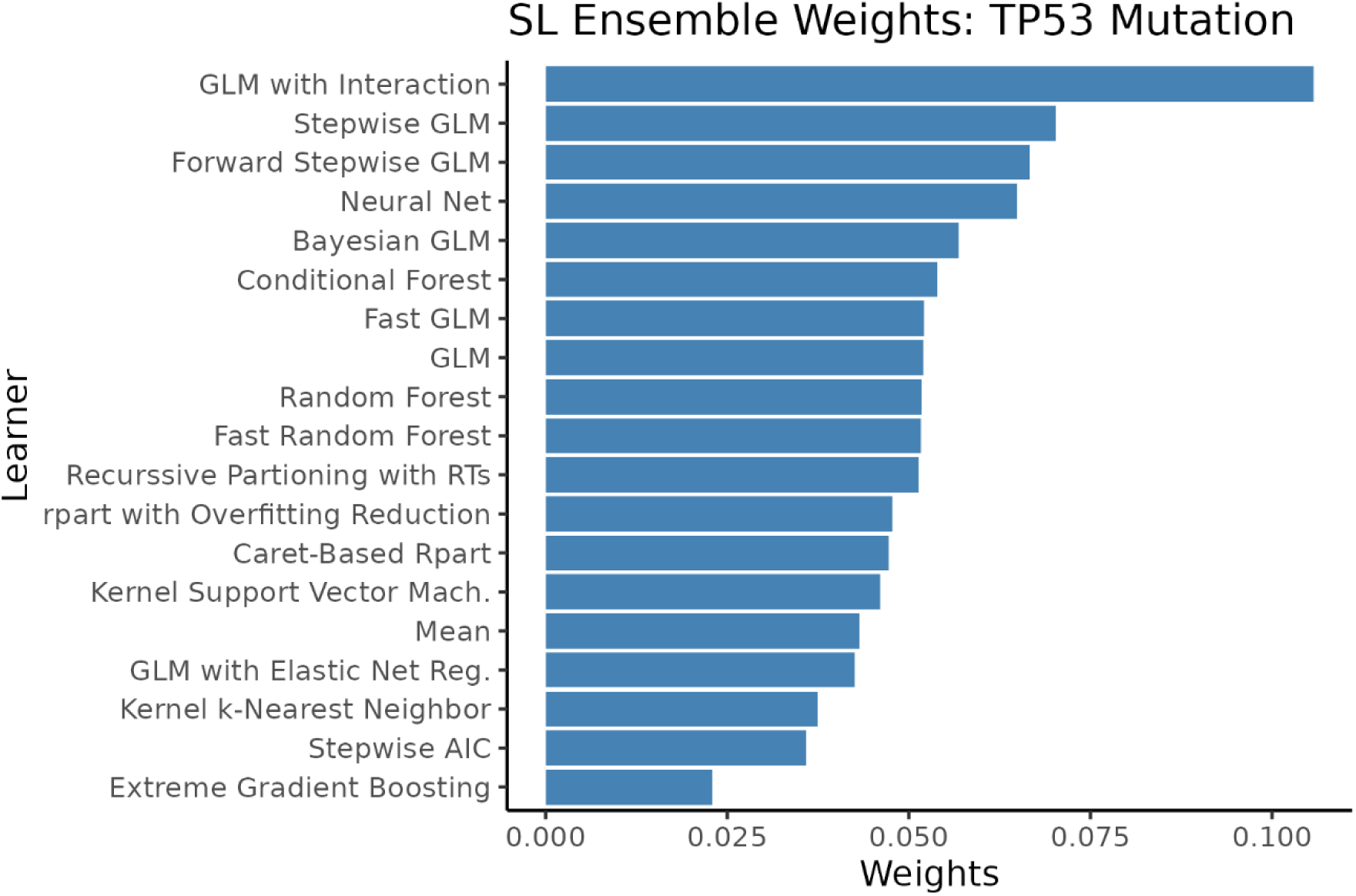
Cross-validated SL ensemble weight averages for binary variable TP53 mutation. *Abbreviations:* Super Learner (SL).

**Figure 4.**
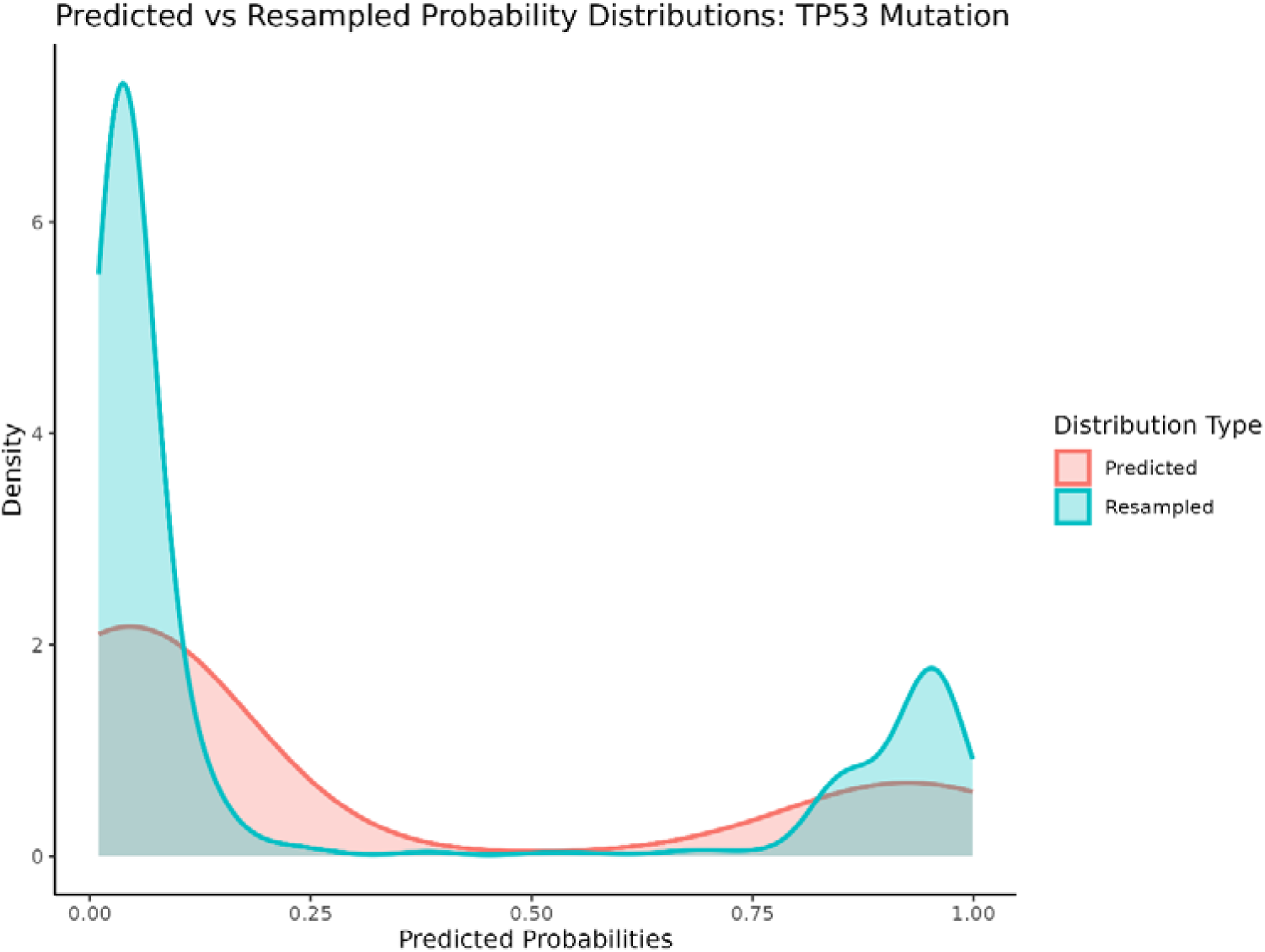
SL-based ensemble’s predicted probability density distribution for TP53 mutation binary variable data with overlapping 1000 resampled density distribution. *Abbreviations:* Super Learner (SL).

**Figure 5.**
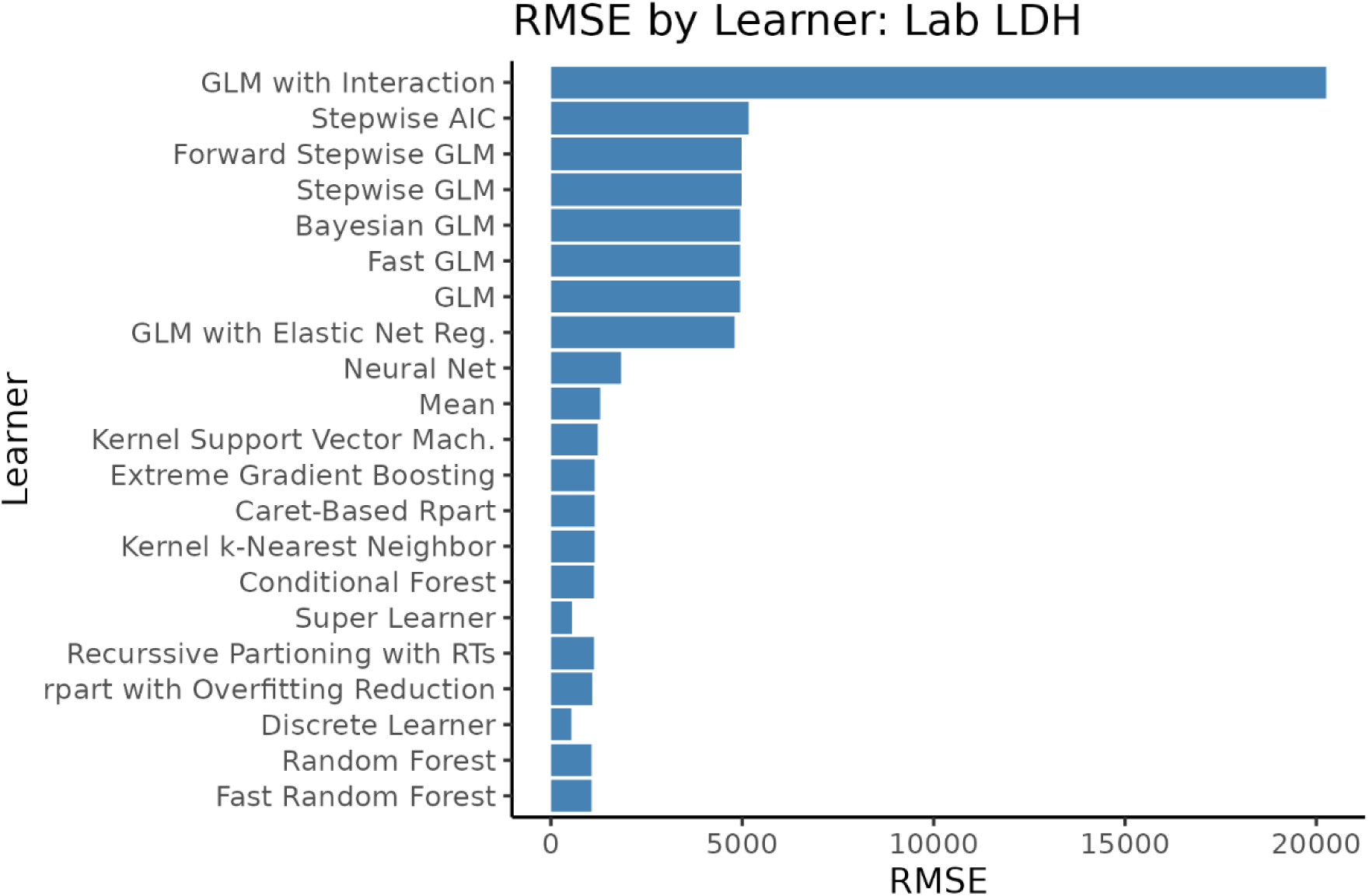
Cross-validated base learner average risks for continuous variable LDH, risk measured as RMSE. *Abbreviations:* Root Mean Square Error (RMSE), Lactate Dehydrogenase (LDH).

**Figure 6.**
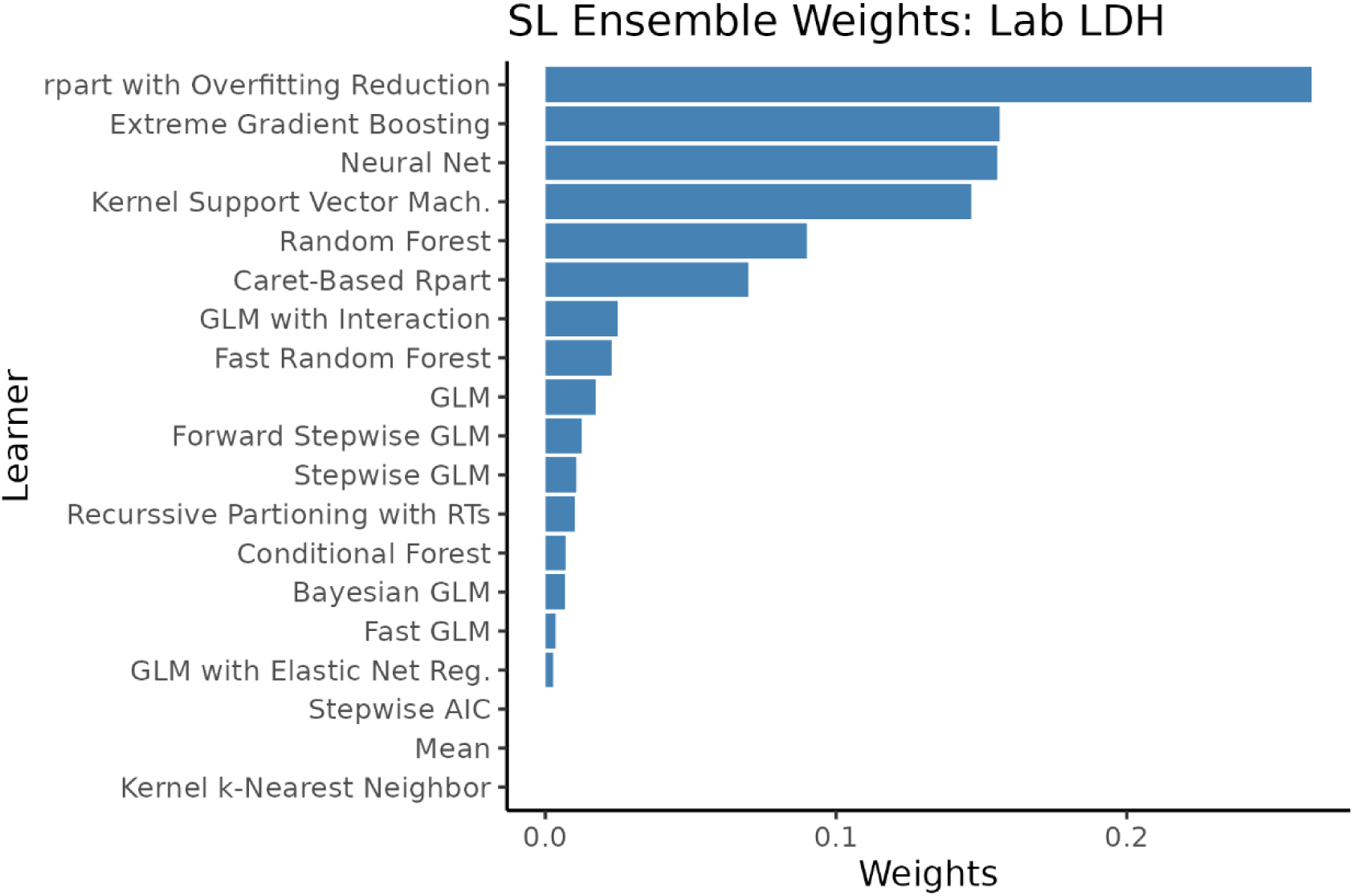
Cross-validated SL-based ensemble average weights for continuous variable LDH. *Abbreviations:* Super Learner (SL), Lactate Dehydrogenase (LDH).

Figure 7 displays the SL ensemble’s predicted values density distribution for LDH and the corresponding density distribution of resampled values based on 1000 resamples, suggesting introduced variation. Subsequent weights indicate moderate to high certainty and relatively small variability of imputations across resamples compared to the observed data’s variation (median 0.81, IQR 0.80-0.83). Lastly, for laboratory measured hemoglobin (SL RMSE: 1.37, MICE RMSE 2.10), platelets (SL RMSE 82.75, MICE RMSE 114.93), and ANC (SL RMSE 2.80, MICE RMSE 4.15) the SL-based ensemble outperformed MICE. Across all 4 continuous variables, this represents an average 24.45% lower RMSE for the SL-based ensemble. The downstream risk stratification comparison between the SL-based imputations and the MICE-based imputations of these risk stratification variables is visualized as survival curve differences in Additional File 9, Figure S8. Using SL-based imputations instead of MICE imputations resulted in 7.78% of patients changing risk classification and a notable increase in separation of median OS days between risk stratifications was observed. Risk group associations with overall survival were consistently stronger under the SL-based imputation. Compared with the favorable group, intermediate risk showed higher hazards under SL (MICE HR 1.21, 95% CI 1.05–1.41; SL HR 1.41, 95% CI 1.21–1.64), and adverse risk was similarly amplified (MICE HR 2.06, 95% CI 1.76–2.41; SL HR 2.36, 95% CI 1.98–2.80), corresponding to an average 25% increase in hazard ratio relative to MICE-derived stratifications. Our computing setup with 48 cores and 188 GB of RAM, with parallelization and a 19-learner library, processed 11 binary targets and a continuous target in approximately 2.5 hours.

**Figure 7.**
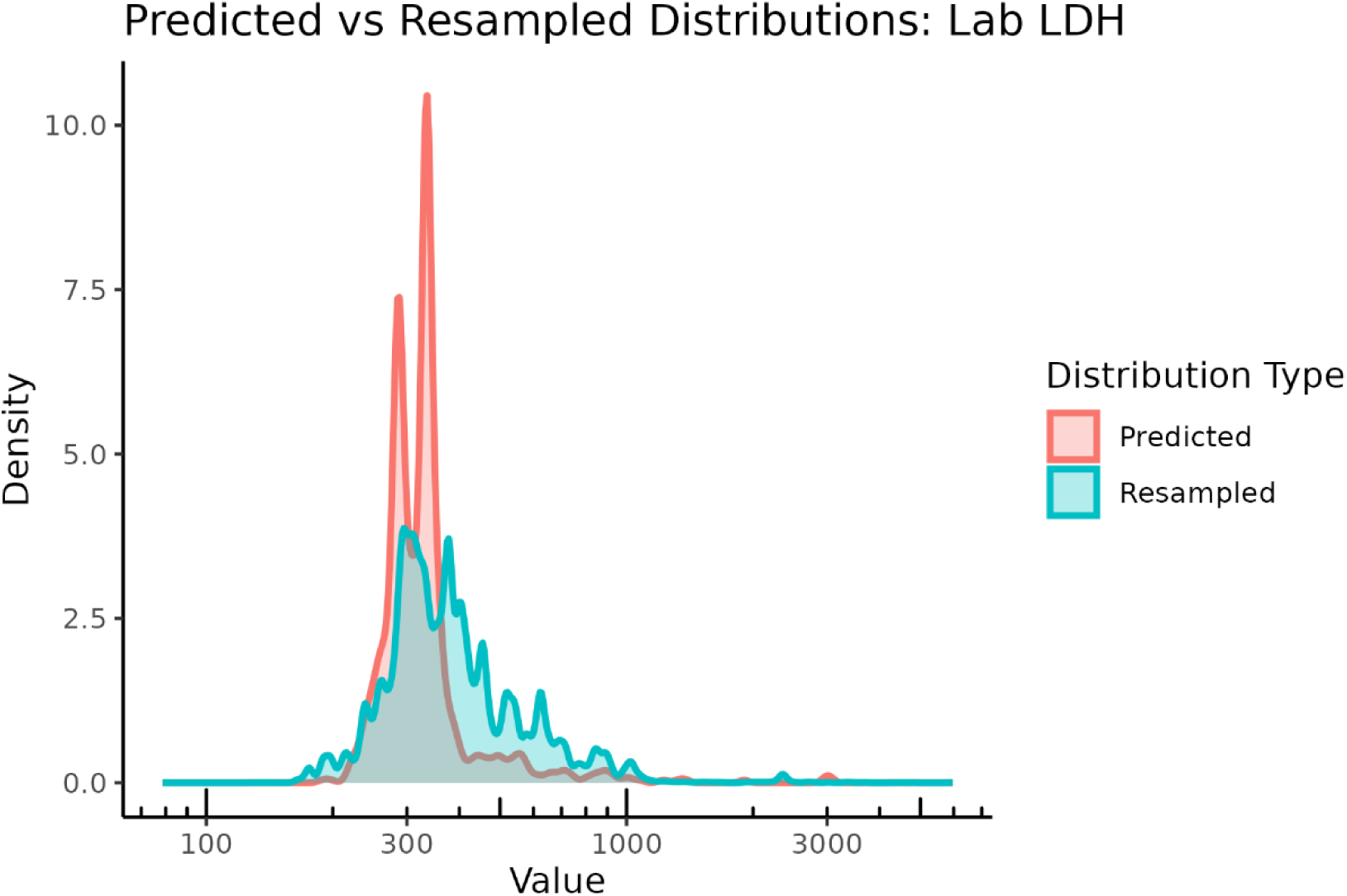
SL-based ensemble’s predicted values density distribution for missing LDH continuous variable data, overlayed with the 1000 resampled density distribution. Figure was log transformed after running the function for interpretability. *Abbreviations:* Lactate Dehydrogenase (LDH).

## Discussion

In this study we evaluated a SL-based ensemble imputation approach for handling missing diagnostic data in a dataset comprised of 1641 newly diagnosed AML patients treated with Ven/Aza or 7+3. Leveraging the SL framework, we demonstrated consistently improved imputation performance over a commonly used statistical method, MICE, specifically for binary genetic variables, even when they had substantial missingness. On average, the SL-based ensemble achieved higher balanced accuracy (89.0% vs 80.8%) and F1 score (96.3% vs 94.3%) compared to MICE across 9 binary genetic features. In focused evaluations of TP53 mutation and LDH laboratory values, the SL-based ensemble approach led to superior or comparable performance to singular algorithms and uncertainty weights. Lastly, in downstream analyses, changes in Kaplan-Meier survival curves and multivariable Cox regression results were noteworthy.

These results underscore the potential of SL-based methods to enhance data integrity in clinical prediction pipelines, especially in high-dimensional electronic health record derived datasets where conventional single-model approaches may struggle. To our knowledge, this study represents one of the first applications of the SL-based ensemble for missing value imputation in a clinical oncology context. Furthermore, we demonstrate the method’s utility by achieving strong performance while introducing novel, observation-level uncertainty estimates which quantify variation, while being delivered through a streamlined, user-friendly function that automates imputation, performance evaluation, and data output.

Prior studies into missing value imputation in clinical cohorts have repeatedly shown that standard approaches such as MICE can yield reasonable accuracy when data are moderately complete and missingness mechanisms approximate MAR, but their performance often deteriorates in high-dimensional or block-missing contexts (7,31). MICE has strong advantages such as being well-documented and easily applied (32). These strengths are evident by the notable abundance of MICE’s application in health research literature, with MICE serving as either the primary imputation method or the benchmark comparison. An earlier 2015 review by Rezvan et al. with narrow search criteria showed the dominance of MICE in imputation methodology in medical research, with 53% (20/38) of research articles reporting methods of imputation using MICE (33). ML-based methods like random forest and k-NN have improved on MICE’s limitations in several ways such as by capturing nonlinear feature relationships (5,8). In the systematic review of 94 ML performance assessing research articles by Alabadla et al. it was found that across most studies k-NN, random forest, and support vector machine methods performed best, these results would agree with our rankings of ML algorithms by AUC and RMSE. This study also noted though the lack of, and need for, computational time reporting and consistency across performance measures to best evaluate overall performance (7). Another study that analyzed high dimensional genetic data compared ML-based approaches to MICE and simple mean imputation, also finding that k-NN and random forest typically performed far better than both of these approaches (34). Such popularity for k-NN, and other algorithms, is explained by a systematic review of 117 research articles by Thomas and Rajabi. The review clarifies that these algorithm’s easy implementation across knowledge domains make them an attractive option. However, these authors also note that despite such popularity there continues to be no single universal solution to the missing data problem (35). While ML-based methods have grown immensely in popularity and application frequency since 2015 (7), in part due to improvements over MICE and simple imputation, as previously mentioned these methods remain relatively constrained by the reliance on a single learner that may have varying performance for different missingness scenarios. Ensemble learning approaches may prove an effective next step to overcome these challenges. While using a more typical ensemble strategy compared to our SL-based strategy, our results would concur with those found by Batra et al. who found that a stacked ensemble strategy proved superior to simple imputation and several singular ML algorithms (12). In a different context than imputation, a 2023 review of 45 articles evaluated ensemble learning approaches in highly researched disease prediction. They concluded that amongst these studies, those that employed the stacking method of ensemble learning performed best compared to other ensemble methods. This work gives us greater confidence in our findings using a SL-based approach (14). Prior literature using super learning for missing data imputation is sparse, however, one study showed that multiple imputation by super learning yielded superior results compared to traditional methods in a comparative analysis. Albeit, this application was limited by synthetic data, not having quantified uncertainty, and a process that did not automatically generate performance measures (15). Lastly, our results extend prior work by demonstrating that a CV-specific stacking ensemble can more effectively integrate varying base learners to handle diverse variable missingness contexts, and when incorporated into a function, can also output measures of uncertainty. Such measures of uncertainty are intended as a practical guardrail for imputation confidence. Uncertainty weights can highlight subject-specific cases where imputations may remain uncertain due to deviating assumptions from cross-validation induced missingness, where uncertainty could be high even when imputation performance across folds is strong.

Despite the promising performance of our SL-based approach, our study has several limitations. First, we employed default SL hyperparameters for all base learners rather than performing extensive tuning. While this decision simplified the pipeline and demonstrated robust gains over MICE by default, it may have missed additional opportunities to further improve performance. Second, the computational burden of training the CV-specific ensemble represents the greatest practical hurdle to routine deployment. However, this cost is justified in settings where imputation accuracy and uncertainty quantification materially impact other modeling tasks in high stakes clinical applications such as risk stratification. As we demonstrated, using the SL-based approach instead of MICE showed noteworthy changes in risk classification, which could have great impacts on clinical treatment planning and health outcomes for patients, especially for aggressive diseases like AML. Regarding the computational time of the whole process, it was also longer than MICE but to quantify uncertainty from MICE using traditional approaches of bootstrapping would likely incur even more computational costs. Lastly, prior to ensemble fitting we perform simple imputation on the top 10 predictors for each target, which may propagate initial error and contribute to cumulative uncertainty. We partially mitigate this through our resampling-based uncertainty weights, observations with high variance can be down-weighted in downstream models. However, such bootstrapping-based uncertainty does not capture within-subject variability and may be susceptible to the amount of missing data and representation of the true missing data pattern in the bootstrap samples. More rigorous initial imputation of predictors, such as using MICE (36), and incorporation of initial subject-specific uncertainty measures in imputation of target variables would further reduce this concern. Conversely, our SL-based imputation function offers several key strengths. By vectorizing imputation across diverse base learners, it provides a nearly fully automated, scalable framework that consistently outperforms single-method approaches in our application in high-dimensional clinical data. Additionally, while we applied the function to AML data for evaluation, there stands no reason it cannot be applied to other datasets. The model’s ability to generate observation-specific uncertainty weights and uncertainty intervals allow end-users to conduct a variety of sensitivity analyses using these values across different settings. The flexibility of the SL framework allows inclusion of new algorithms as they emerge, ensuring that the pipeline can adapt to evolving patterns of data complexity and missingness. Finally, in line with identified gaps in performance assessment in the literature (37) and combined with descriptive statistics of missingness, the SL-based imputation function’s performance output allows a user to well understand key aspects of MVI including computational time and common performance measures.

## Conclusions

In summary, our work demonstrates that a SL-based imputation function can significantly improve the accuracy and robustness of missing-value estimation in a high-dimensional AML real-world dataset, outperforming the standard MICE approach for most of our target variables while providing observation-level uncertainty metrics. These findings support our central hypothesis that ensemble strategies yield more reliable imputation in complex clinical data. In future deployments we hope to add automated and customized hyperparameter optimization, integrate automatic support for multinomial and ordinal targets, and further validate its utility in diverse missing data formats and contexts. Future research would benefit from evaluating this function among diverse disease areas, across a greater range of variables, its impact on optimizing risk stratification strategies, and in more extensive simulation and validation studies that assess uncertainty calibration. Ultimately, by combining improved performance over traditional methods and flexibility, ensemble learning-derived imputation can enhance downstream modeling by managing missingness’ threat to validity.

## Abbreviations

|  |  |
| --- | --- |
| AML | Acute myeloid leukemia |
| AUC | Area under the curve |
| CU | University of Colorado |
| CYT | Cytogenetics |
| k-NN | k-Nearest Neighbor |
| LAB | Laboratory values |
| LDH | Lactate dehydrogenase |
| MAR | Missing at random |
| MAD | Median absolute deviation |
| MCAR | Missing completely at random |
| MICE | Multiple imputation by chained equations |
| ML | Machine learning |
| MVI | Missing value imputation |
| NNLS | Non-negative least squares |
| RMSE | Root mean square error |
| RWC | Real World Cohort |
| RWD | Real world data |
| SL | Super Learner |
| Ven/Aza | Venetoclax plus azacitidine |
| $\hat{p}_i$ | Predicted probability |
| $\tau$ | Optimal threshold |
| $B_i$ | Bootstrap samples |
| $p_{\square_i}$ | Mean predicted value |

## Declarations

### Ethics approval and consent to participate

This study was conducted in accordance with the principles of the Declaration of Helsinki. This study was reviewed and approved under the 2018 Requirements of the Federal Policy for the Protection of Human Subjects by the Colorado Multiple Institutional Review Board (COMIRB), located at the University of Colorado Anschutz Medical Campus, Aurora, Colorado, United States. Ethical approval was granted for the study, and a Waiver of Informed Consent and a Health Insurance Portability and Accountability Act (HIPAA) Waiver of Authorization were granted by COMIRB on the basis that all criteria for waiving consent and HIPAA authorization were met. The Flatiron Health data that supported the findings of this study were originated by and are the property of Flatiron Health, Inc.”

### Consent for publication

Not applicable.

### Availability of data and materials

The individual patient-level datasets analyzed during the current study are not publicly available due to data privacy laws but the processed data are available from the corresponding author on reasonable request. The Flatiron Health data that support the findings of this study are available from Flatiron Health Inc., but restrictions apply to the availability of these data, which were used under license for th current study, and so are not publicly available. Flatiron Health data has restriction prohibiting the authors from making the datasets publicly available.

Requests for data sharing by license or by permission for the specific purpose of replicating results in this manuscript can be submitted to. Lastly, in the supplementary material, the SL-based imputation function is accompanied by an HTML-based illustration that provides a concise, step-by-step walkthrough that enables users to quickly understand its purpose, inputs, outputs, and performance metrics, and to apply the function to their own datasets. This HTML-based illustration serves as a tutorial which includes the full function code and applies the method to synthetic data. A simple R package to apply the function is forthcoming.

### Author’s contributions

TZ contributed via conception and design of the work, analysis of the data, interpretation of the data, creation of the function applied, and drafting and revising the work. LW contributed via design of the work and acquisition of the data. JD, JC, KS, FM, and UK contributed via acquisition and management of the data. JR contributed via acquisition and management of data and internal review of the manuscript. NI contributed via conception and design of the work, acquisition and interpretation of the data, and revising the manuscript. All authors read and approved the final manuscript.

## Supporting information

SL-Based imputation Function Illustration

## Data Availability

The raw, individual patient data are protected and not available due to data privacy laws. The processed data are available at reasonable requests to the corresponding author. The Flatiron Health data that supported the findings of this study originated by and are the property of Flatiron Health, Inc., which has restrictions prohibiting the authors from making the data set publicly available. Requests for data sharing by license or by permission for the specific purpose of replicating results in this manuscript can be submitted to. Lastly, upon publication, the SL-based imputation function will be accompanied by an HTML-based illustration that provides a concise, step-by-step walkthrough that enables users to quickly understand its purpose, inputs, outputs, and performance metrics, and to apply the function to their own datasets.

## Acknowledgements

We give additional acknowledgement and great gratitude to the internal reviewers of RefinedScience and external reviewers of Flatiron Health.

## Additional Files

**Additional file 1, png:** Figure S1. Molecular Landscape Comparison of CU and RWC. CU and RWC merged cohort’s OncoPlot, representing AML phenotypic and genotypic features per-patient. +ve refers to the number of patients with positive features and the % refers to that positive feature proportion. Light grey regions represent missing data. *Abbreviations:* University of Colorado (CU), Real World Cohort (RWC).

**Additional file 2, docx:** Table S1. Default, and used, SL algorithm library with descriptions. All SL learner algorithms used in our default library within the SL-based imputation function.

*Abbreviations:* Super Learner (SL).

**Additional file 3, png:** Figure S2. Merged Cohort Variable Missingness. CU and RWC merged cohort’s variable missingness ratios. *Abbreviations:* University of Colorado (CU), Real World Cohort (RWC).

**Additional file 4, png:** Figure S3. Merged Cohort Missingness Heatmap – Cytogenetic Testing. CU and RWC merged cohort’s missingness heatmap for all observations and binary cytogenetics variables. *Abbreviations:* University of Colorado (CU), Real World Cohort (RWC), Cytogenetics (CYT).

**Additional file 5, png:** Figure S4. Merged Cohort Missingness Heatmap – Mutations. CU and RWC merged cohort’s missingness heatmap for all observations and binary mutation variables. *Abbreviations:* University of Colorado (CU), Real World Cohort (RWC).

**Additional file 6, png:** Figure S5. Merged Cohort Missingness Heatmap – Lab Values. CU and RWC merged cohort’s missingness heatmap for all observations and continuous laboratory values variables. *Abbreviations:* University of Colorado (CU), Real World Cohort (RWC), Laboratory Values (LAB).

**Additional file 7, png:** Figure S6. Top 10 Predictors for TP53 Mutation. Top 10 predictors for the TP53 mutation binary variable from the merged CU and RWC cohorts. *Abbreviations:* University of Colorado (CU), Real World Cohort (RWC).

**Additional file 8, png:** Figure S7. Top 10 Predictors for LDH Lab Value. Top 10 predictors for the LDH continuous variable from the merged CU cohort and RWC. *Abbreviations:* Lactate Dehydrogenase (LDH), University of Colorado (CU), Real World Cohort (RWC).

**Additional file 9, png:** Figure S8. Risk stratification-stratified Kaplan Meier OS curves comparing the SL-based ensemble’s imputation of risk stratification variables and the MICE-based imputation of risk stratification variables. Note, imputations for this additional analysis were re-run on updated data, reflecting 6 new observations. *Abbreviations:* Overall Survival (OS), Multiple Imputations by Chained Equations (MICE), Super Learner (SL), Log-Rank test (LR), Tarone-Ware test (TW), Fleming-Harrington early-weighted test (FH:e), Fleming-Harrington late-weighted test (FH:l).

**Additional file 10, html:** HTML-based illustration that contains the core MVI function code in its most recent version as well as serving as a tutorial of the function’s arguments, outputs, and how to apply it in an example using synthetic data.

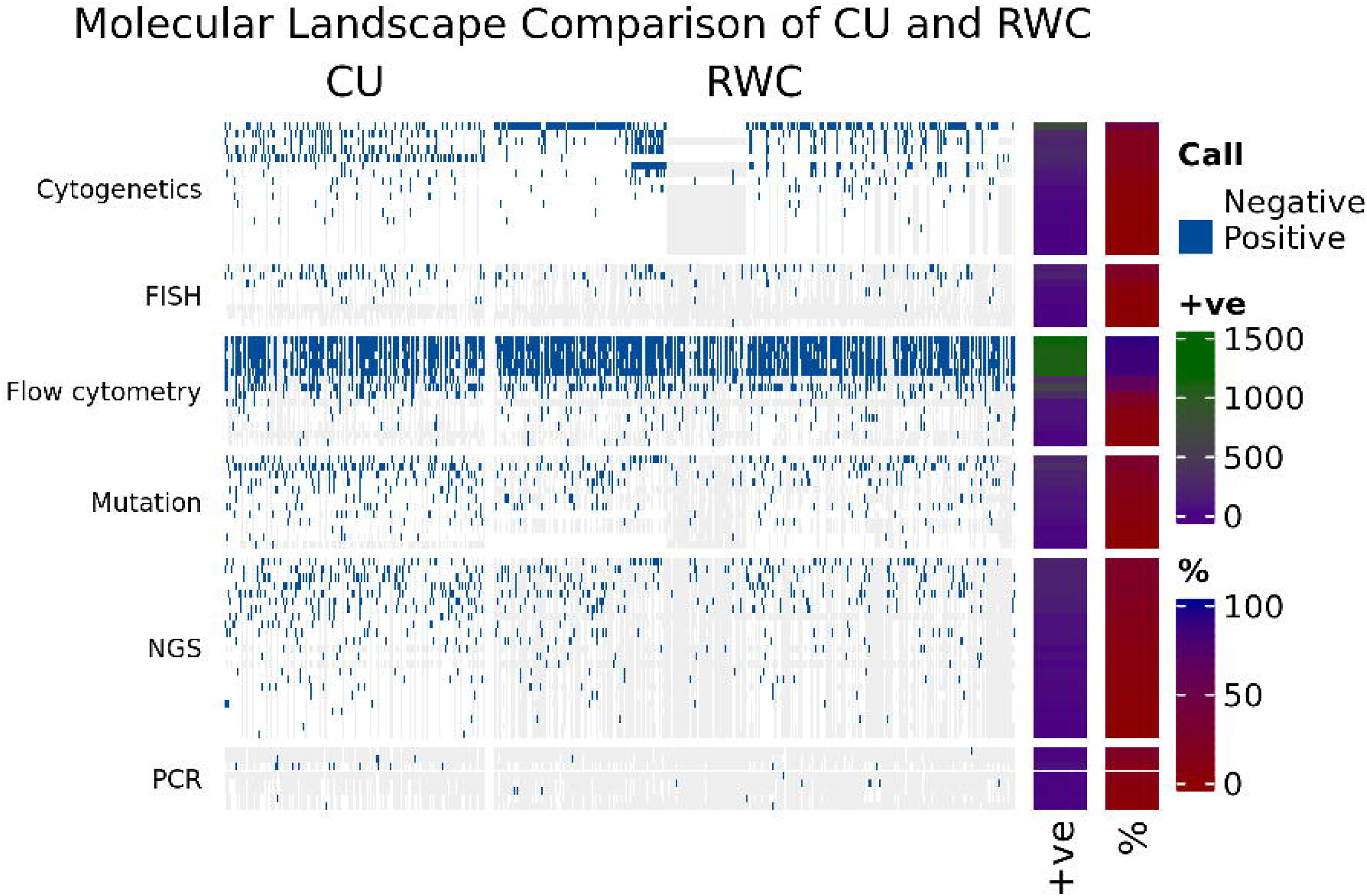

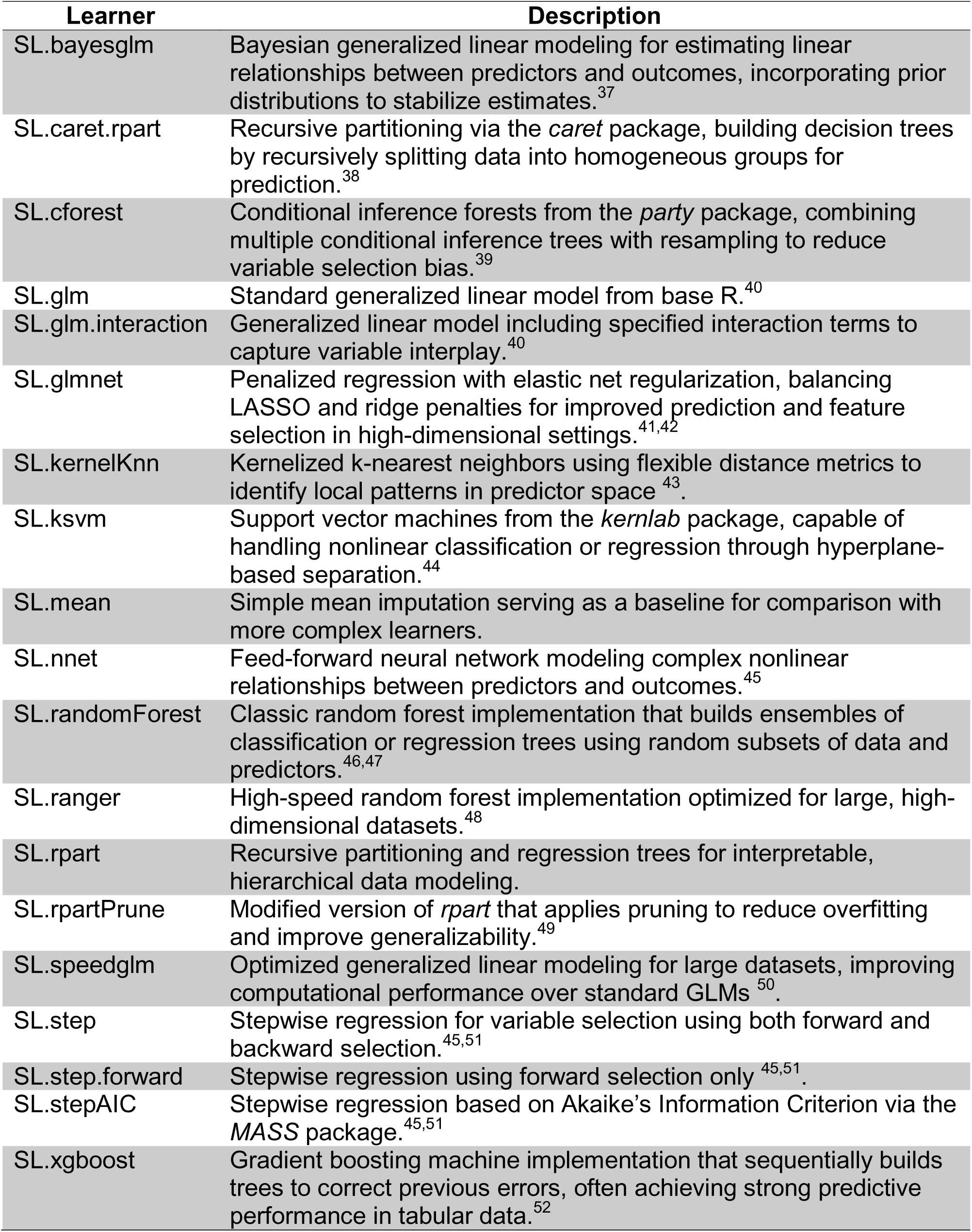

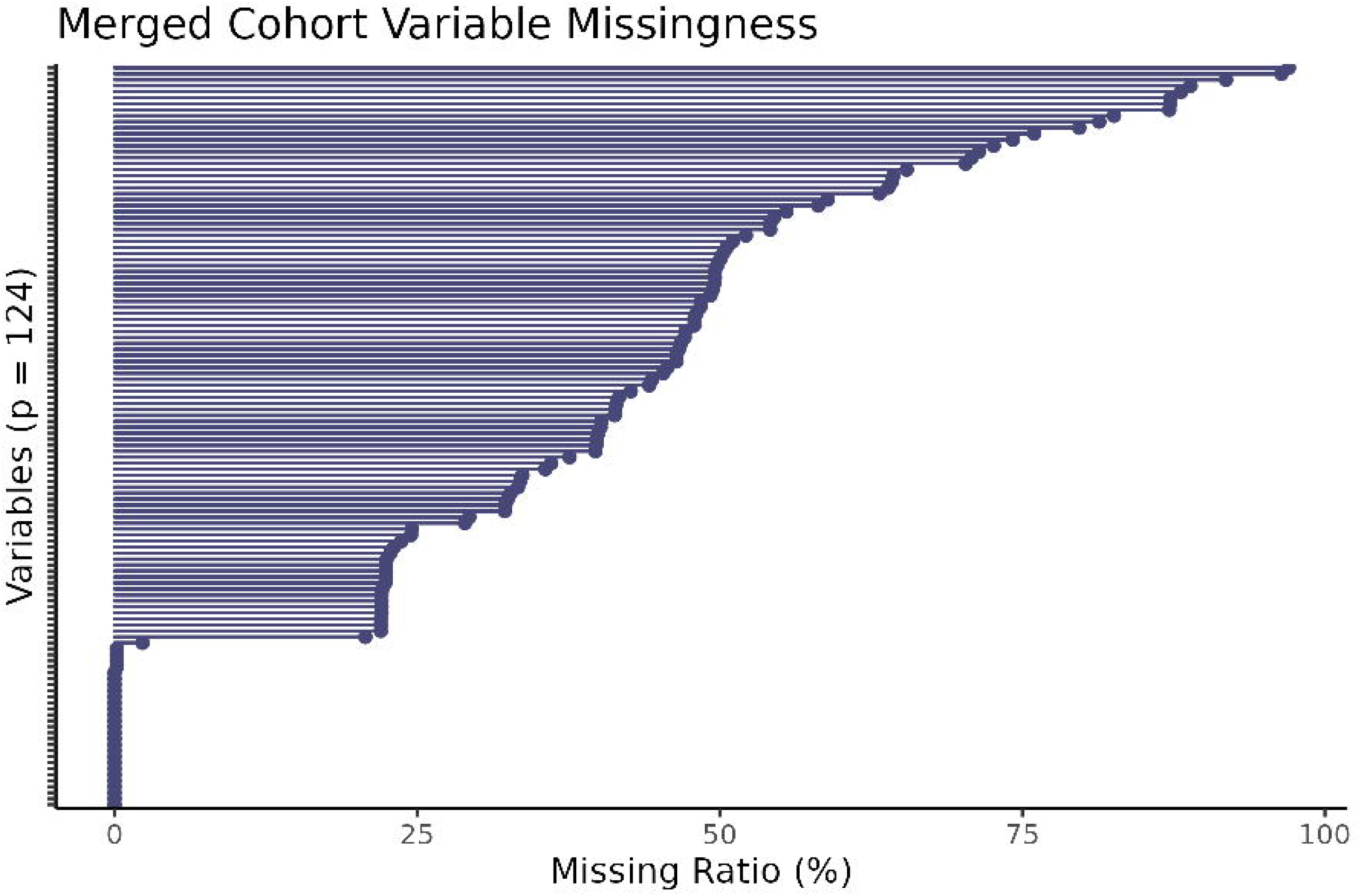

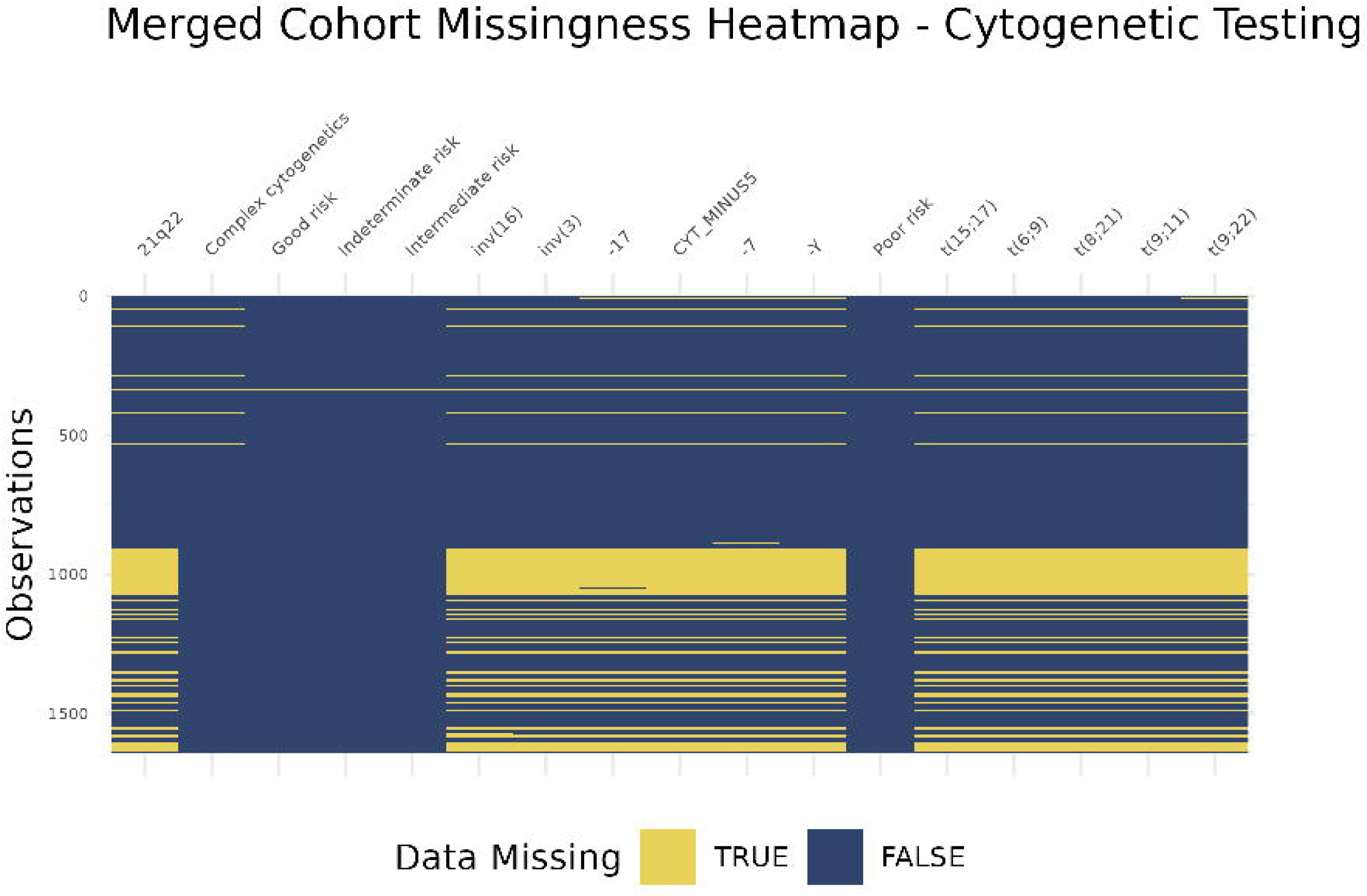

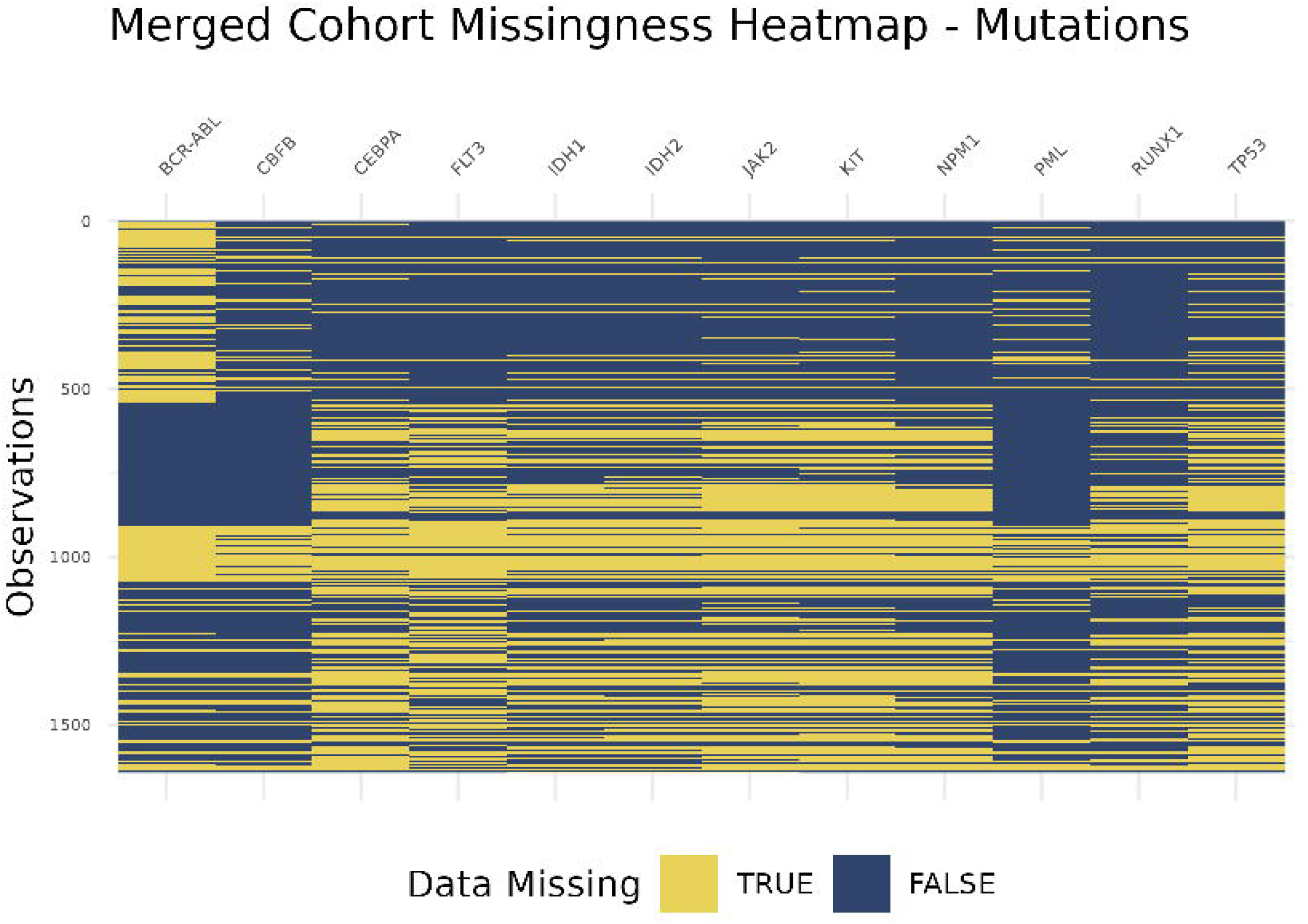

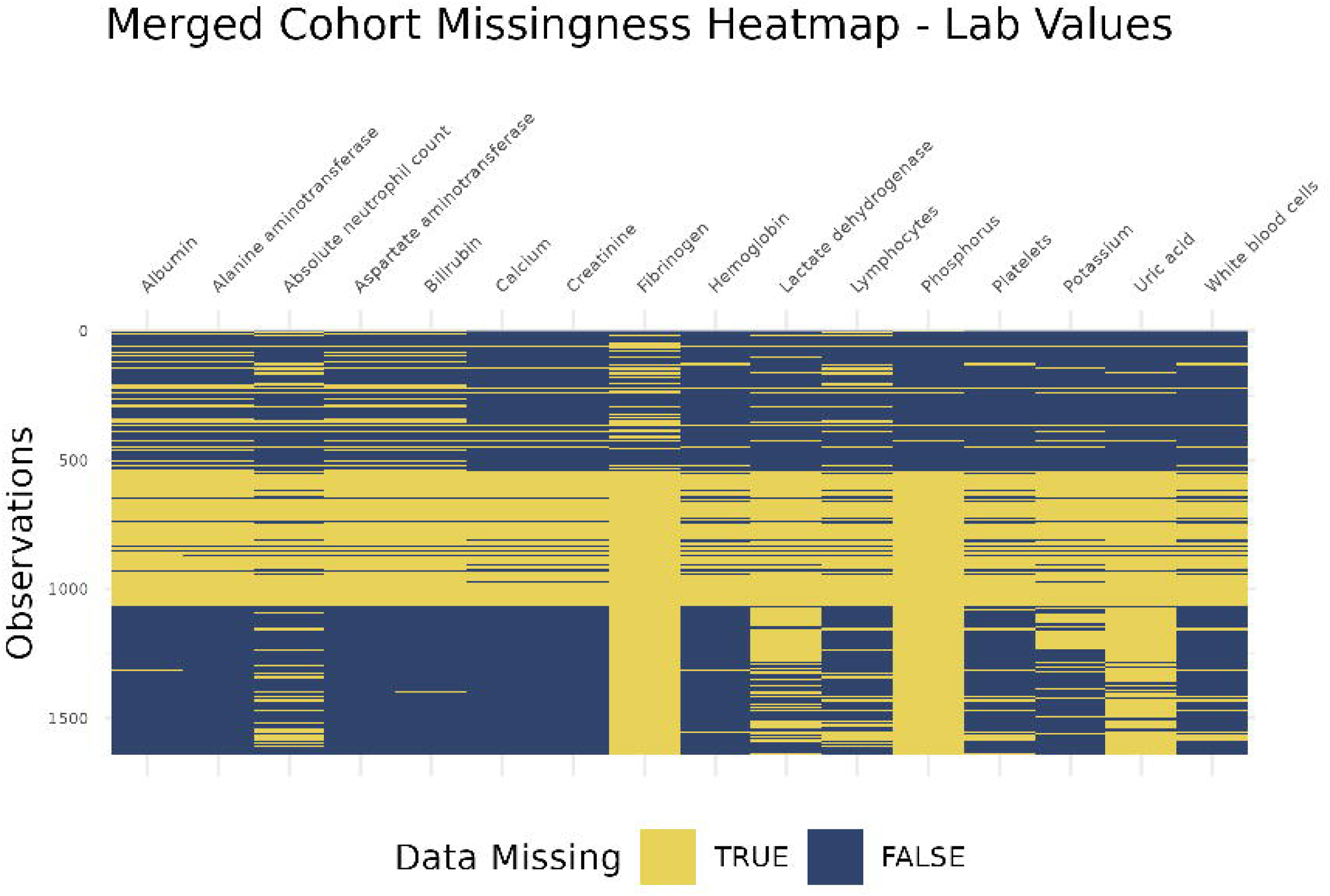

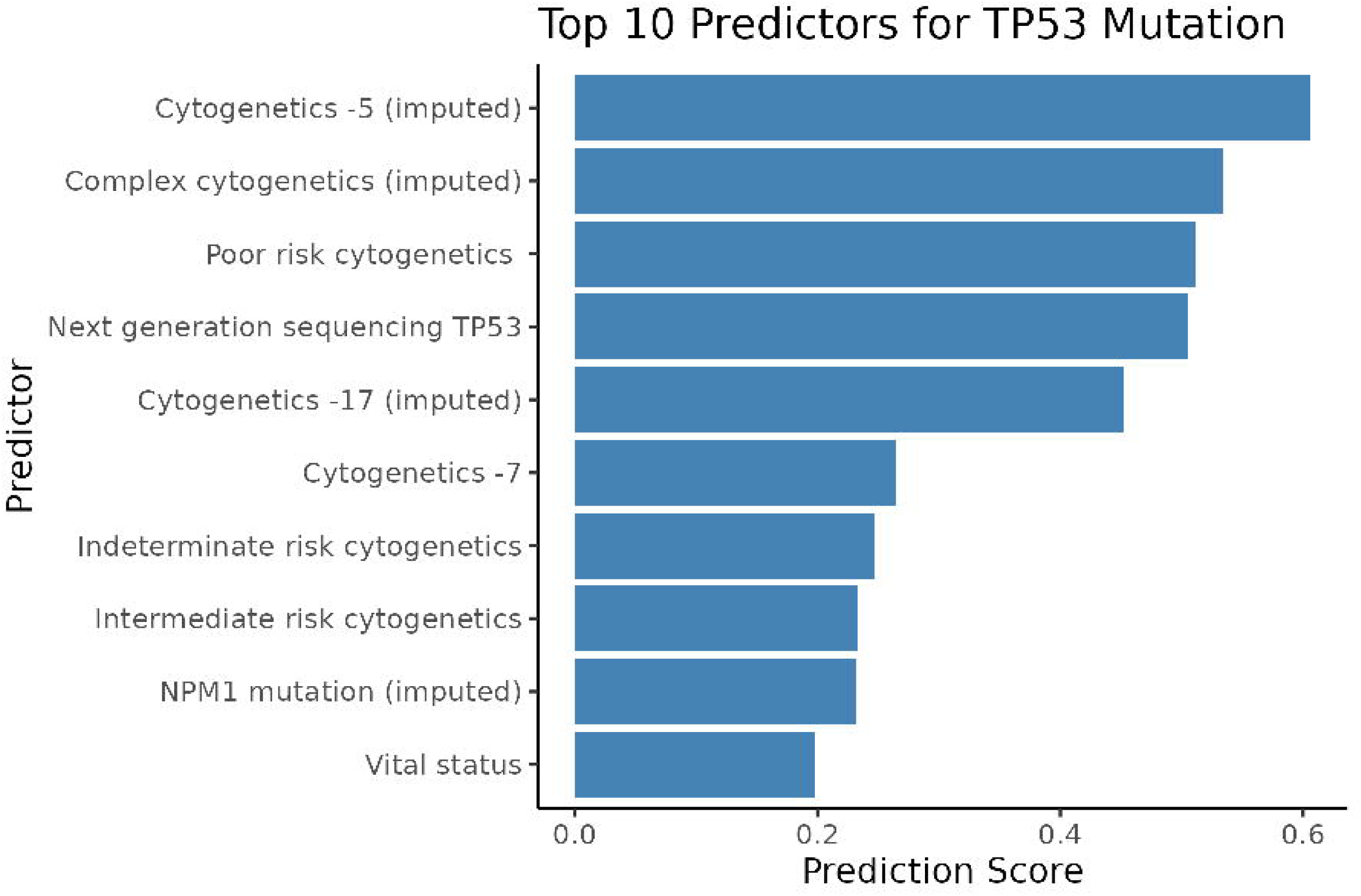

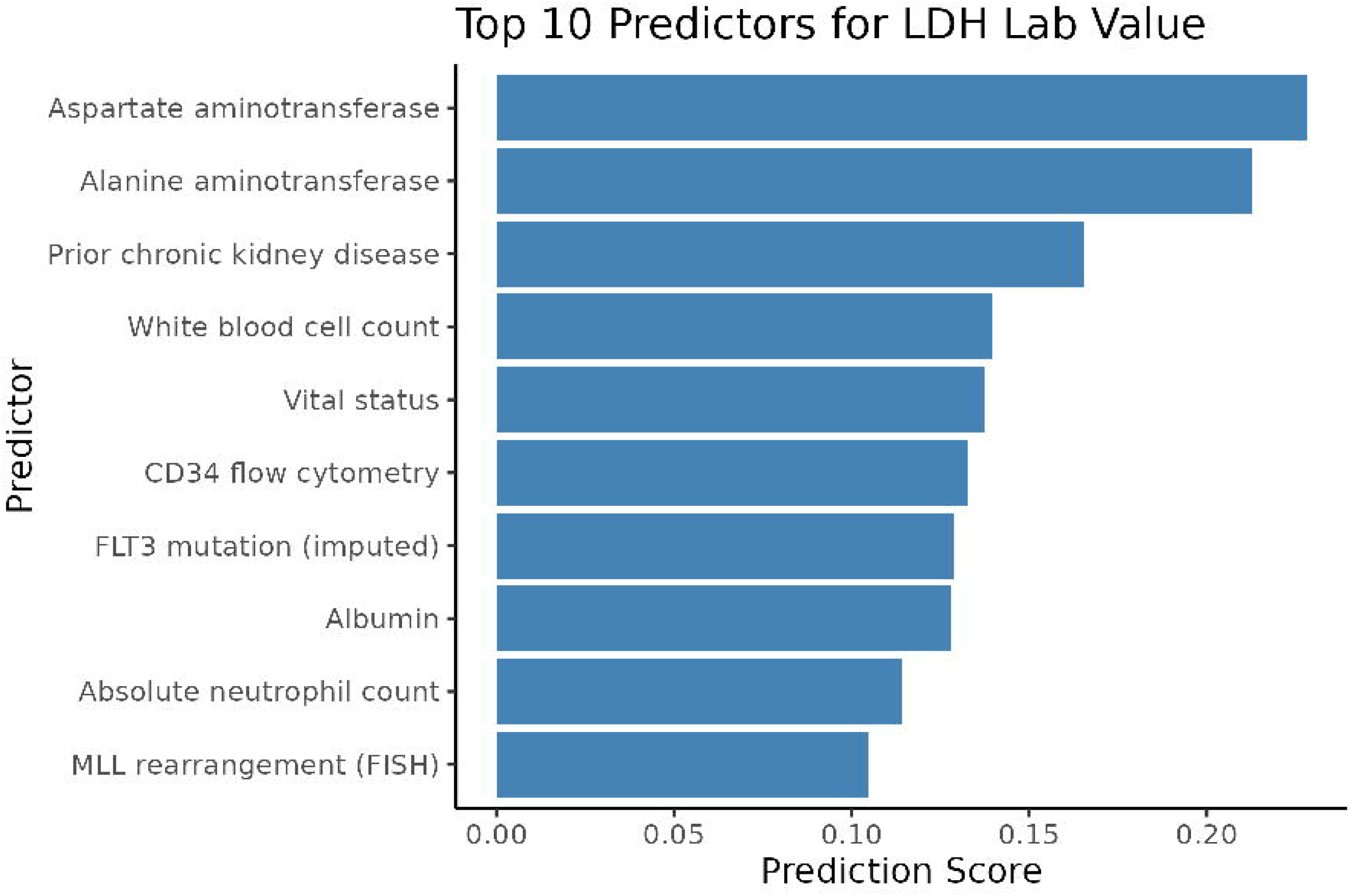

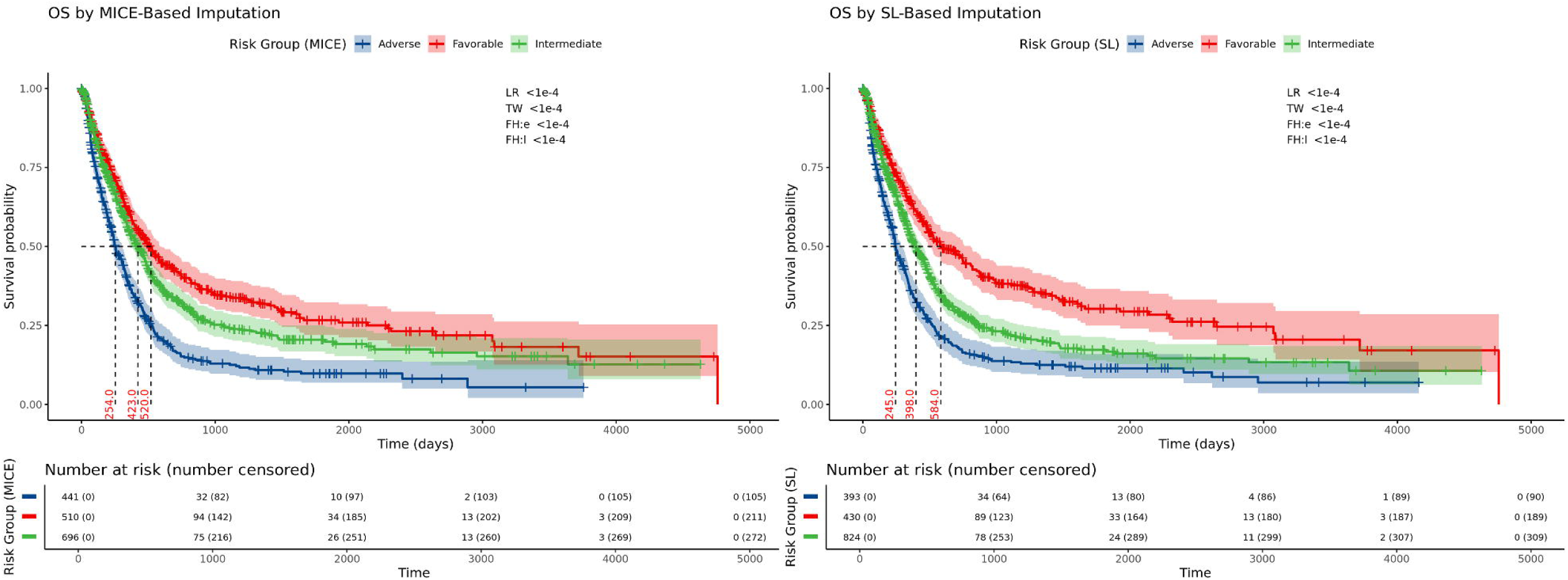

