## Supplementary material for "A Comparative Analysis in a Clinical Cohort: Multiple Imputation by Chained Equations and a Novel Super Learner-Based Imputation Approach": SL-Based imputation Function Illustration: AF10 SL-Based-Imputation-Function-Illustration.html

An illustration for the development of a novel SL-based missing value imputation function for simulated acute myeloid leukemia patient data


Code 

- Show All Code
- Hide All Code

### An illustration for the development of a novel SL-based missing value imputation function for simulated acute myeloid leukemia patient data

###### Tony Zbysinski, PhD


#### 01/23/2026

### Overview

This HTML report demonstrates the implementation and functionality of
a novel Super Learner (SL)-based missing value imputation (MVI) function
developed in conjunction with the associated manuscript. A synthetic
dataset consisting of 1000 subjects and 100 features was generated to
support this demonstration, including simulated missingness relevant to
real-world data. It is important to note that this dataset is strictly
illustrative and not intended for clinical interpretation; due to
inherent limitations in simulating realistic missing data mechanisms,
results should be interpreted with caution. The report is organized into
six sections: beginning with required R package loading, followed by a
description of the SL-based MVI function, generation of the synthetic
dataset, visualization and evaluation of function outputs for both
binary and continuous variables, and concluding with a demonstration of
full vector-based imputation. Together, these sections provide a
comprehensive and reproducible guide for understanding the workflow and
performance of the SL-based imputation function.

### Necessary packages, parallelization, and conflict management

We first must load the necessary R packages to run the function, we
additionally will manage some function conflicts and setup
parallelization to drastically decrease computational time.

```
options(warn = -1)
knitr::opts_chunk$set(
  error = FALSE,
  results = 'hide',
  warning = FALSE,
  message = FALSE,
  cache = TRUE,
  autodep = TRUE,
  cache.lazy = FALSE,
  dpi = 150, 
  fig.retina=1)

### Libraries
library(bigrquery)
library(tidyverse)
library(naniar)
library(SuperLearner)
library(viridis)
library(knitr)
library(mice)
library(corrr)
library(furrr)
library(glmnet)
library(randomForest)
library(gam)
library(e1071)
library(arm)
library(party)
library(gbm)
library(LogicReg)
library(polspline)
library(ipred)
library(earth)
library(caret)
library(conflicted)
library(parallel)
library(doParallel)
library(xgboost)
library(cvAUC)
library(pROC)
library(speedglm)
library(glmnet)
library(KernelKnn)
library(kernlab)
library(MASS)
library(ranger)
library(SIS)
library(parallel)
library(future.apply)
library(kableExtra)

### Conflict management
conflict_prefer("select","dplyr")
conflict_prefer("filter", "dplyr")

### Parallelization
## Parallelization for SuperLearner
## The commented out parts are outdated, have moved onto the "snow" method of parallelization further below
# # Get number of cores and set it
# num_cores = RhpcBLASctl::get_num_cores()
# options(mc.cores = num_cores)
# 
# # Set seed for across cores
# set.seed(123, "L'Ecuyer-CMRG")
# 
# ## Parellelization for bootstrapping
# future::plan("multisession")

if (!exists("sl_cluster", inherits = TRUE) || !inherits(sl_cluster, "cluster")) {
  n <- max(1, parallel::detectCores() - 1)
  sl_cluster <- parallel::makeCluster(n)
  parallel::clusterSetRNGStream(sl_cluster, 1)

  invisible(parallel::clusterEvalQ(sl_cluster, {
    library(SuperLearner)
  }))
}

options(future.globals.maxSize = 2*1024^3) # 2GB for carrying of the larger objects in continuous variable imputation

## Fixing figure sizing for illustration
theme_set(theme_classic(base_size = 18))
```

### Description of functions

This function implements a SL-based imputation strategy for
high-dimensional datasets with missing binary and continuous variables.
It operates in two modes or separate functions: a single-variable mode
for imputation and performance evaluation of one variable at a time, and
a vectorized mode that sequentially imputes a list of variables,
allowing earlier imputations to inform later ones. The latter function
incorporates the former and can handle single variables, meaning that
the only function that needs to be used is the vectorized form. The
approach uses a stacked ensemble model to optimize estimation accuracy
and generate observation-specific uncertainty measures for downstream
analyses. The function is tailored for clinical and real-world data
contexts with complex missingness patterns and serves as both a robust
imputation engine and a diagnostic tool for evaluating imputation
quality.

The function requires a clean dataset containing only binary and
continuous variables. Variables such as non-numeric categories, unknown
flags, data variables, or upper/lower reference bounds must be removed
prior to use. A unique identifier variable is encouraged, as while the
inner function’s workings uses a row counter to track and organize the
data, a unique identifier can be specified that allows the user to more
easily merge newly imputed data back with other datasets. The remaining
variables need to be converted to a numeric class of binary or
continuous description. The main arguments are:

1. `target_vars`: Character vector of variables to be
   imputed (ordered by least to most missingness or importance).
2. `df_input`: Input dataset with required
   format.
3. `learners`: Character vector of SL base learner
   wrappers.
4. `screeners`: Character vector of optional screener
   wrappers.
5. `id_var`: The unique identifier variable in the
   dataset.
6. `n_top_predictors`: The number of top predictors to be
   used to estimate the missing data, default is 10 with a recommended
   5-15.

Computational requirements are substantial. In our use case, we used
48 cores and 189 GB of memory. Even with parallelization and a ~19
learner SL library, binary variable imputations took ~10 minutes each,
and continuous variables ~20 minutes. Continuous variables likely take
considerably longer given observation-level resampling for uncertainty.
We recommend limiting imputations to no more than 10 variables at a time
(maximum 3 continuous variable at a time) under similar computing
conditions.

1. `missrank()` — ranks a data frame’s variables from
   least to most missing.
2. `rs_mvifunc()` — single-variable SL–based imputation
   used in `rs_multimvifunc()`.
3. `rs_multimvifunc()` — multi-variable SL–based
   imputation for a specified vector of target variable, or it can handle
   one variable at a time.

```
### Function to get and extract vector of missing variables in order of least to most missing
missrank <- function(df) {
  miss_ratios <- colMeans(is.na(df))
  sorted_vars <- sort(miss_ratios, decreasing = FALSE)
  names(sorted_vars)
}

### Single Missing Variable Imputation Function
## Specifying the function
rs_mvifunc <- function(target_var, df_input, 
                       learners = c("SL.bayesglm", "SL.caret.rpart", "SL.cforest",
                       "SL.glm", "SL.glm.interaction",
                       "SL.glmnet", "SL.kernelKnn", "SL.ksvm", 
                       "SL.mean", "SL.nnet", "SL.randomForest", 
                       "SL.ranger", "SL.rpart", "SL.rpartPrune", "SL.speedglm",
                       "SL.step", "SL.step.forward", "SL.stepAIC",
                       "SL.xgboost"), #default
                       screeners = c("All"), #default, no additional screeners
                       var_type = c("binary", "continuous"),
                       id_var = NULL, #default none
                       n_top_predictors = 10) { #default 10
  
  ## Take variable and makes sure it runs the right pipeline for the datatype
  var_type <- match.arg(var_type)
  
  ## Generate a .rowid if it doesn't already exist
  if (!".rowid" %in% names(df_input)) {
    df_input <- dplyr::mutate(df_input, .rowid = dplyr::row_number())
  }
  
  if (!is.null(id_var)) {
    if (!id_var %in% names(df_input)) {
        stop(sprintf("id_var '%s' not found in df_input.", id_var))
    }
  }
  
  ## Function that takes input learners and screeners, creating all combinations
  make_SL_library <- function(learners, screeners) {
      library_list <- list()
      
      library_list <- c(library_list, as.list(learners))
      
      for (screener in screeners) {
        for (learner in learners) {
          library_list <- append(library_list, list(c(learner, screener)))
        }
      }
      
      return(library_list)
    }

  ## Actually creating the SL.library
  SL.library <- make_SL_library(learners, screeners)
  
  ## Adding a naming map for user friendliness when reading plots downstream
  learner_name_map <- c(
    "SL.bayesglm"        = "Bayesian GLM",
    "SL.bartMachine"     = "Bayesian Additive RTs",
    "SL.biglasso"        = "Big Lasso",
    "SL.caret"           = "Caret (Generic)",
    "SL.caret.rpart"     = "Caret-Based Rpart",
    "SL.cforest"         = "Conditional Forest",
    "SL.earth"           = "Multivariate Adaptive Reg. Splines",
    "SL.gam"             = "Generalized Additive Model",
    "SL.gbm"             = "Gradient Boosting Mach.",
    "SL.glm"             = "GLM",
    "SL.glm.interaction" = "GLM with Interaction",
    "SL.glmnet"          = "GLM with Elastic Net Reg.",
    "SL.ipredbagg"       = "Bagged GLMs",
    "SL.kernelKnn"       = "Kernel k-Nearest Neighbor",
    "SL.knn"             = "k-Nearest Neighbor",
    "SL.ksvm"            = "Kernel Support Vector Mach.",
    "SL.lda"             = "Linear Discriminant Analysis",
    "SL.leekasso"        = "Regularized Linear Reg.",
    "SL.lm"              = "Linear Model",
    "SL.loess"           = "Local Polynomial Reg.",
    "SL.logreg"          = "Logistic Reg.",
    "SL.mean"            = "Mean",
    "SL.nnet"            = "Neural Net",
    "SL.nnls"            = "Non-Negative LS Reg.",
    "SL.polymars"        = "Piecewise Polynomial Function Reg.",
    "SL.qda"             = "Quadratic Discriminant Analysis",
    "SL.randomForest"    = "Random Forest",
    "SL.ranger"          = "Fast Random Forest",
    "SL.ridge"           = "Reg. with Ridge Penalty",
    "SL.rpart"           = "Recurssive Partioning with RTs",
    "SL.rpartPrune"      = "rpart with Overfitting Reduction",
    "SL.speedglm"        = "Fast GLM",
    "SL.speedlm"         = "Fast LM",
    "SL.step"            = "Stepwise GLM",
    "SL.step.forward"    = "Forward Stepwise GLM",
    "SL.step.interaction"= "Stepwise GLM (Interaction)",
    "SL.stepAIC"         = "Stepwise AIC",
    "SL.svm"             = "Support Vector Machine",
    "SL.xgboost"         = "Extreme Gradient Boosting"
  )
  
  ## 1. Interim-processing
  # Dropping iteratively made variables that are irrelevant to prediction
  drop_vars <- names(df_input)[
    names(df_input) == ".rowid" |
    (!is.null(id_var) & names(df_input) == id_var) |
    str_detect(names(df_input), "(weight|CI_lower|CI_upper|lower_imp|upper_imp)$")
  ]
  
  df_filtered <- df_input[, !(names(df_input) %in% drop_vars)]

  # Getting missing ratios of all variables in the dataset for predictor selection
  df_missratios <- map_dbl(df_filtered, ~ mean(is.na(.x)))
  
  # Isolating just the potential predictors
  allnames <- setdiff(names(df_missratios), target_var)

  # Flag for high missingness
  high_missing_flag <- df_missratios[[target_var]] > 0.8
  if (high_missing_flag) {
    message(paste("Warning:", target_var, "has >80% missingness."))
  } else {
    message(paste(target_var, "does not have >80% missingness."))
  }

  # Grabbing correlation scores and var names for predictor selection
  corr_scores <- map_dbl(allnames, ~ cor(df_input[[target_var]], df_input[[.x]], use = "pairwise.complete.obs"))
  names(corr_scores) <- allnames

  # Formula balancing correlation with missingness for predictor selection
  predscores <- abs(corr_scores) * (1 - df_missratios[names(corr_scores)])
  
  # Grabbing top predictors
  ordered <- sort(predscores, decreasing = TRUE)
  k <- min(n_top_predictors, length(ordered))
  topkpreds <- names(ordered)[seq_len(k)]
  toppreds <- setdiff(topkpreds, target_var)

  # Plotting top predictors and each one's score
  top10_df <- data.frame(Predictor = toppreds, Score = predscores[toppreds])
  top10_plot <- ggplot(top10_df, aes(x = reorder(Predictor, Score), y = Score)) +
    geom_col(fill = "steelblue") +
    coord_flip() +
    theme_classic() +
    labs(title = paste("Top 10 Predictors for", target_var), x = "Predictor", y = "Prediction Score")

  ## 2. Impute top predictors via simple imputation (numeric: median, binary: mode)
  df_imp <- df_input

  for (var in toppreds) {
    var_data <- df_input[[var]]
    unique_vals <- unique(var_data[!is.na(var_data)])

    if (is.numeric(var_data) && length(unique_vals) == 2) {
      fill_value <- as.numeric(names(sort(table(var_data), decreasing = TRUE))[1])
    } else if (is.numeric(var_data)) {
      fill_value <- median(var_data, na.rm = TRUE)
    } else {
      fill_value <- names(sort(table(var_data), decreasing = TRUE))[1]
    }

    var_data[is.na(var_data)] <- fill_value
    df_imp[[var]] <- var_data
  }

  ## 3. Subset data (predictors and target var) for SL
  sldata <- df_imp %>%
    select(all_of(toppreds), all_of(target_var)) %>%
    filter(!is.na(.data[[target_var]]))

  ## 4a. Binary imputation pipeline
  if (var_type == "binary") {
    # Check for extreme class imbalance because it'll probably fail the SL process if too imbalanced
    class_dist <- table(sldata[[target_var]]) / nrow(sldata)
    max_class_prop <- max(class_dist)

    ## Imbalanced sub-pipeline: uses simple (mode) imputation
    targclass <- sldata[[target_var]]
    n_pos  <- sum(targclass == 1, na.rm = TRUE)
    n_neg  <- sum(targclass == 0, na.rm = TRUE)
    
    if (max_class_prop >= 0.925 || min(n_pos, n_neg) <= 20) {
      mode_val <- as.numeric(names(which.max(table(sldata[[target_var]]))))
      mode_weight <- max_class_prop
      mode_class <- mode_val

      class_imbalance <- paste0(
        "Class imbalance detected (",
        round(mode_weight * 100, 2),
        "% of class ", mode_class,
        ") — used mode imputation."
      )
      
      # Create and make numeric new imputed variables value's, weight equal to the proportion
      imp_final <- df_input %>%
        filter(is.na(.data[[target_var]])) %>%
        transmute(
          .rowid,
          !!paste0(target_var, "simimp") := as.numeric(mode_val),
          !!paste0(target_var, "weight") := as.numeric(mode_weight)
        )

      # Take observed variable's values and rename
      obs_final <- df_input %>%
        filter(!is.na(.data[[target_var]])) %>%
        transmute(
          .rowid,
          !!paste0(target_var, "simimp") := .data[[target_var]],
          !!paste0(target_var, "weight") := 1
        )

      # Merge original observed and imputed target variables into one
      full_column <- bind_rows(obs_final, imp_final)

      df_augmented <- df_input %>%
        left_join(full_column, by = ".rowid") %>% 
        select(-target_var)

      # Getting just the target var's results
      df_merged <- df_input %>%                     
        dplyr::left_join(full_column, by = ".rowid")
      
      imp_cols <- grep(paste0("^", target_var, "(imp|simimp)$"), names(df_merged), value = TRUE)
      aux_cols <- grep("(weight|CI_lower|CI_upper|lower_imp|upper_imp)$", names(df_merged), value = TRUE)
      
      keep_cols <- c(".rowid",
                     if (!is.null(id_var)) id_var,
                     target_var,
                     imp_cols,
                     aux_cols)
      keep_cols <- unique(keep_cols[keep_cols %in% names(df_merged)])
      
      var_data <- df_merged %>% dplyr::select(dplyr::all_of(keep_cols))

      # Return list for simple (mode) imputation for highly imbalanced classes
      return(list(
        top10_plot = top10_plot,
        class_imbalance = class_imbalance,
        imputed_data = imp_final,
        updated_data = df_augmented,
        var_data = var_data,
        high_missing = high_missing_flag
      ))
    }

    # Recalling top predictors, isolating that data, creating a prediction matrix
    toppreds_test <- toppreds
    df_test <- df_input[, c(target_var, toppreds_test)]
    pred_matrix <- make.predictorMatrix(df_test)
    pred_matrix[,] <- 0
    pred_matrix[target_var, toppreds_test] <- 1

    # Setting seed for making observed and masked data for MICE comparison
    observed <- which(!is.na(df_test[[target_var]]))
    mask <- sample(observed, round(0.3 * length(observed)))

    truevals <- df_test[[target_var]][mask]
    df_test_masked <- df_test
    df_test_masked[[target_var]][mask] <- NA

    # MICE comparison test and confusion matrix for performance measurement
    mice_time <- system.time({
      imp_test <- mice(df_test_masked, m = 5, predictorMatrix = pred_matrix)
      completed_test <- complete(imp_test)
      imputedvals <- completed_test[[target_var]][mask]
    })

    mice_confmatrix <- confusionMatrix(factor(imputedvals), factor(truevals))

    # Calculating F1 for imbalance class cases as an additional performance metric
    mice_recall <- caret::recall(as.factor(truevals), as.factor(imputedvals))
    mice_precision <- caret::precision(as.factor(truevals), 
                                       as.factor(imputedvals))
    mice_f1 <- 2 * (mice_precision * mice_recall) / 
      (mice_precision + mice_recall + 1e-10)
    #this is a spot for future work, confusionMatrix has a hidden F1 that can be pulled instead of recalculating
    
    # Setting special seed for parallelization of SL and setting library
    test.library <- SL.library

    # Cross-validation loop (of the process) for SL performance measurement
    slcv <- CV.SuperLearner(
      Y = sldata[[target_var]],
      X = sldata[, toppreds],
      family = binomial(),
      parallel = sl_cluster,
      SL.library = test.library,
      method = "method.AUC",
      verbose = TRUE
    )

    # Extracting raw learner names and isolating refined labels
    learners_raw <- slcv$libraryNames
    learners_clean <- gsub("(_.*)?$", "", learners_raw)
    
    # Making learners names better with name map
    pretty_labels <- ifelse(learners_clean %in% names(learner_name_map),
                      learner_name_map[learners_clean],
                      learners_raw)  #fallback to raw names
    names(pretty_labels) <- learners_raw

    # Extracting true and predicted probability values from the cross-validated process
    y_true <- slcv$Y
    y_pred_prob <- slcv$SL.predict
    ydisc_prob <- slcv$discreteSL.predict
    
    # Creating optimal threshold for classification of predicted probabilities, default uses AUC to optimize but uses F1 if <95% and >90% class imbalance (very rare)
    obs_rate <- mean(y_true)
    threshold_metric <- if (obs_rate < 0.1 || obs_rate > 0.9) "F1" else "AUC"

    opt_threshold <- if (threshold_metric == "F1") {
      scores <- sapply(seq(0.01, 0.99, by = 0.01), function(th) {
        y_pred_class <- ifelse(y_pred_prob >= th, 1, 0)
        cm <- confusionMatrix(factor(y_pred_class), factor(y_true))
        2 * (cm$byClass["Precision"] * cm$byClass["Recall"]) / 
          (cm$byClass["Precision"] + cm$byClass["Recall"] + 1e-10)
      })
      seq(0.01, 0.99, by = 0.01)[which.max(scores)]
    } else {
      scores <- sapply(seq(0.01, 0.99, by = 0.01), function(th) {
        y_pred_class <- ifelse(y_pred_prob >= th, 1, 0)
        auc(roc(y_true, y_pred_class))
      })
      seq(0.01, 0.99, by = 0.01)[which.max(scores)]
    }

    # Repeating for discrete single best algorithm
    obs_rate <- mean(y_true)
    threshold_metric <- if (obs_rate < 0.1 || obs_rate > 0.9) "F1" else "AUC"

    discopt_threshold <- if (threshold_metric == "F1") {
      scores <- sapply(seq(0.01, 0.99, by = 0.01), function(th) {
        y_pred_class <- ifelse(ydisc_prob >= th, 1, 0)
        cm <- confusionMatrix(factor(y_pred_class), factor(y_true))
        2 * (cm$byClass["Precision"] * cm$byClass["Recall"]) / 
          (cm$byClass["Precision"] + cm$byClass["Recall"] + 1e-10)
      })
      seq(0.01, 0.99, by = 0.01)[which.max(scores)]
    } else {
      scores <- sapply(seq(0.01, 0.99, by = 0.01), function(th) {
        y_pred_class <- ifelse(ydisc_prob >= th, 1, 0)
        auc(roc(y_true, y_pred_class))
      })
      seq(0.01, 0.99, by = 0.01)[which.max(scores)]
    }
    
    # Applying optimal threshold
    y_pred_class <- ifelse(y_pred_prob >= opt_threshold, 1, 0)
    ydisc_pred_class <- ifelse(ydisc_prob >= discopt_threshold, 1, 0)
    
    # Calculating confusion matrix, AUC, and other performance measures 
    sl_conf <- confusionMatrix(factor(y_pred_class), factor(y_true))
    sl_table <- sl_conf$table
    
    sldisc_conf <- confusionMatrix(factor(ydisc_pred_class), factor(y_true))
    
    sl_recall <- caret::recall(as.factor(y_true), as.factor(y_pred_class))
    sl_precision <- caret::precision(as.factor(y_true), as.factor(y_pred_class))
    sl_f1 <- 2 * (sl_precision * sl_recall) / (sl_precision + sl_recall + 1e-10)

    # Plotting binary AUC performance of each base learner and SL
    slcv_plot <- plot(slcv) +
      scale_x_discrete(labels = pretty_labels) +
      labs(title = paste("Cross-Validated Base Learner Risks: ", target_var)) +
      xlab("Learner") +
      ylab("Risk (AUC)") +
      theme_classic()

    # Moving onto normal SL application for prediction
    sl_time <- system.time ({  
      slfull <- snowSuperLearner(
        Y = sldata[[target_var]],
        X = sldata[, toppreds],
        family = binomial(),
        cluster = sl_cluster,
        SL.library = test.library,
        method = "method.AUC",
        verbose = TRUE
      )
    })
      
    # Subsetting only the missing data for prediction and predicting
    miss_data <- df_imp %>% filter(is.na(.data[[target_var]])) %>% select(all_of(toppreds))
    miss_preds <- predict(slfull, newdata = miss_data, onlySL = TRUE)

    # Number of bootstrap samples for resampling for balanced accuracy CIs
    B <- 1000

    miss_p <- as.numeric(miss_preds$pred)
    
    # Balanced Accuracy
    balanced_acc <- function(y_true, y_pred) {
      cm <- confusionMatrix(factor(y_pred), factor(y_true))
      sens <- cm$byClass["Sensitivity"]
      spec <- cm$byClass["Specificity"]
      mean(c(sens, spec), na.rm = TRUE)
    }
    
    # Bootstrap loop
    boot_bal_acc <- replicate(B, {
      boot_idx <- sample(seq_along(y_true), replace = TRUE)
      boot_probs <- y_pred_prob[boot_idx]
      boot_labels <- y_true[boot_idx]
      boot_preds <- ifelse(boot_probs >= opt_threshold, 1, 0)
      balanced_acc(boot_labels, boot_preds)
    })
    
    # Calculate CI and mean for Balanced Accuracy
    ba_ci_lower <- quantile(boot_bal_acc, 0.025, na.rm = TRUE)
    ba_ci_upper <- quantile(boot_bal_acc, 0.975, na.rm = TRUE)
    ba_mean <- mean(boot_bal_acc, na.rm = TRUE)
    
    resample_ba <- c(ba_mean, ba_ci_lower, ba_ci_upper)
    
    # Observation-level bootstrapped predictions
    sim_mat <- replicate(B, {
      boot_idx <- sample(seq_along(miss_p), replace = TRUE)
      miss_p[boot_idx]
    })
    
    lower_ci <- apply(sim_mat, 1, quantile, probs = 0.025, na.rm = TRUE)
    upper_ci <- apply(sim_mat, 1, quantile, probs = 0.975, na.rm = TRUE)
    
    ci_lower_class <- ifelse(lower_ci >= opt_threshold, 1, 0)
    ci_upper_class <- ifelse(upper_ci >= opt_threshold, 1, 0)
    
    # Lower and upper imputation weights retired because the formula doesn't work like that, need a new one if desired but unlikely to be relevant
    # ci_lower_weight <- abs(lower_ci - opt_threshold) / 
    #   max(opt_threshold, 1 - opt_threshold)
    # ci_upper_weight <- abs(upper_ci - opt_threshold) / 
    #   max(opt_threshold, 1 - opt_threshold)
    
    # Transpose and pivot to long format
    sim_long <- as.data.frame(t(sim_mat)) %>%
      tibble::rownames_to_column(var = "Observation") %>%
      tidyr::pivot_longer(
        cols = starts_with("V"),
        names_to = "BootstrapSample",
        values_to = "value"
      ) %>%
      mutate(type = "Resampled")
    
    # Original predictions
    obs_long <- tibble(
      Observation = as.character(seq_along(y_pred_prob)),
      BootstrapSample = "Original",
      value = y_pred_prob,
      type = "Predicted"
    )
    
    combined_data <- bind_rows(sim_long, obs_long)
    
    # Density Plot of predicted probabilities and resampled distribution
    pred_dist_plot <- ggplot(combined_data, aes(x = value, color = type, fill = type)) +
      geom_density(alpha = 0.3, size = 1, adjust = 1.5) +
      theme_classic() +
      labs(
        title = paste0("Predicted vs Resampled Probability Distributions: ", target_var),
        x = "Predicted Probabilities",
        y = "Density",
        color = "Distribution Type",
        fill = "Distribution Type"
      )

    # Extracting learner coefficients (weights) and plotting
    slcv_coefs <- colMeans(slcv$coef)

    weights_df <- tibble(
      Learner = names(slcv_coefs),
      Weight = as.numeric(slcv_coefs)
    )
    
    weight_plot <- ggplot(weights_df,
                          aes(x = reorder(Learner, Weight), y = Weight)) +
      scale_x_discrete(labels = pretty_labels) +
      geom_col(fill = "steelblue") +
      coord_flip() +
      theme_classic() +
      labs(title = paste("SL Ensemble Weights: ", target_var),
           x = "Learner", y = "Weights")

    # Taking imputed data and creating variables for merging
    imp_final <- miss_data %>%
      mutate(.rowid = df_input %>% 
               filter(is.na(.data[[target_var]])) %>% 
               pull(.rowid), !!paste0(target_var, "imp") := 
               as.numeric(ifelse(miss_preds$pred >= opt_threshold, 1, 0)),
             # Calculating weights as distance from optimal threshold
             !!paste0(target_var, "weight") := 
               as.numeric((abs(miss_preds$pred-opt_threshold))/max(
                 opt_threshold, 1-opt_threshold)),
             !!paste0(target_var, "CI_lower_imp") := ci_lower_class,
             !!paste0(target_var, "CI_upper_imp") := ci_upper_class) %>% 
             # !!paste0(target_var, "CI_lower_weight") := ci_lower_weight,
             # !!paste0(target_var, "CI_upper_weight") := ci_upper_weight) %>%
      select(.rowid, !!paste0(target_var, "imp"), ends_with("weight"), 
             ends_with("lower_imp"), ends_with("upper_imp"))

    # Taking observed data and making compatible for merging
    obs_final <- df_input %>%
      filter(!is.na(.data[[target_var]])) %>%
      transmute(
        .rowid,
        !!paste0(target_var, "imp") := .data[[target_var]],
        !!paste0(target_var, "weight") := 1
      )

    # Merging and consolidating the observed and imputed data
    full_column <- bind_rows(obs_final, imp_final)
    
    df_augmented <- df_input %>%
      left_join(full_column, by = ".rowid") %>% 
      select(-target_var)

    # Getting just the target var's results
    df_merged <- df_input %>%                     
      dplyr::left_join(full_column, by = ".rowid")
    
    imp_cols <- grep(paste0("^", target_var, "(imp|simimp)$"), names(df_merged), value = TRUE)
    aux_cols <- grep("(weight|CI_lower|CI_upper|lower_imp|upper_imp)$", names(df_merged), value = TRUE)
    
    keep_cols <- c(".rowid",
                   if (!is.null(id_var)) id_var,
                   target_var,
                   imp_cols,
                   aux_cols)
    keep_cols <- unique(keep_cols[keep_cols %in% names(df_merged)])
    
    var_data <- df_merged %>% dplyr::select(dplyr::all_of(keep_cols))
    
    # Returning list of diagnostics and data for user
    return(list(
      top10_plot = top10_plot,
      mice_confmatrix = mice_confmatrix,
      mice_f1 = mice_f1,
      slcv_plot = slcv_plot,
      sl_confusion = sl_conf,
      sldisc_confusion = sldisc_conf,
      sl_resample_ba = resample_ba,
      sl_f1 = sl_f1,
      sl_table = sl_table,
      sl_weights_plot = weight_plot,
      opt_threshold = opt_threshold,
      prediction_distribution = pred_dist_plot,
      updated_data = df_augmented,
      var_data = var_data,
      high_missing = high_missing_flag,
      mice_time_sec = mice_time["elapsed"],
      sl_time_sec = sl_time["elapsed"]
    ))
    ## 4b. Continuous/numeric pipeline
  } else {

    # Scaling step for target variable since large continuous scale is troublesome
    y_obs <- sldata[[target_var]]
    min_y <- min(y_obs, na.rm = TRUE)
    max_y <- max(y_obs, na.rm = TRUE)
    range_y <- max_y - min_y
    
    # Avoid division by zero if constant target, unlikely needed safeguard
    if(range_y == 0) {
      y_scaled <- y_obs
    } else {
      y_scaled <- (y_obs - min_y) / range_y
    }

    # Replace target variable in sldata with scaled version
    sldata[[target_var]] <- y_scaled

    # Build prediction matrix for MICE (no scaling)
    toppreds_test <- toppreds
    df_test <- df_input[, c(target_var, toppreds_test)]
    pred_matrix <- make.predictorMatrix(df_test)
    pred_matrix[,] <- 0
    pred_matrix[target_var, toppreds_test] <- 1

    # Setting seed and developing masked and holdout data
    observed <- which(!is.na(df_test[[target_var]]))
    mask <- sample(observed, round(0.3 * length(observed)))
    truevals <- df_test[[target_var]][mask]
    df_test_masked <- df_test
    df_test_masked[[target_var]][mask] <- NA

    # Independent MICE run
    mice_time <- system.time({
      imp_test <- mice(df_test_masked, m = 5, predictorMatrix = pred_matrix)
      completed_test <- complete(imp_test)
      imputedvals <- completed_test[[target_var]][mask]
    })

    # Mice performance measure
    mice_rmse <- sqrt(mean((imputedvals - truevals)^2))

    # Setting special seed and assigning library
    test.library <- SL.library

    # Running cross validation loop (of the process) for performance assessment
    slcv <- CV.SuperLearner(
      Y = sldata[[target_var]],
      X = sldata[, toppreds],
      family = gaussian(),
      parallel = sl_cluster,
      SL.library = test.library,
      method = "method.NNLS",
      verbose = TRUE
    )

    # Getting learner names and reassinging with name map, this could probably be made better
    learners_raw <- slcv$libraryNames
    learners_clean <- gsub("(_.*)?$", "", learners_raw)
    pretty_labels <- ifelse(learners_clean %in% names(learner_name_map),
                            learner_name_map[learners_clean], learners_raw)
    names(pretty_labels) <- learners_raw

    V <- length(slcv$AllSL)
    learner_names <- names(slcv$AllSL$`1`$cvRisk)
    cv_risks_matrix <- matrix(NA, nrow = V, ncol = length(learner_names))
    colnames(cv_risks_matrix) <- learner_names
    for (i in seq_len(V)) {
      cv_risks_matrix[i, ] <- slcv$AllSL[[as.character(i)]]$cvRisk
    }

    # Calculating RMSE for performance for all learners
    mean_mse <- colMeans(cv_risks_matrix, na.rm = TRUE)
    mean_rmse <- sqrt(mean_mse) * range_y  # convert RMSE back to original scale

    y_true <- slcv$Y * range_y + min_y
    sly_pred <- slcv$SL.predict * range_y + min_y
    sldiscy_pred <- slcv$discreteSL.predict * range_y + min_y

    # Manually getting RMSE for SL and single-best learner as the package does not auto-calculate RMSE
    sl_rmse <- sqrt(mean((y_true - sly_pred)^2, na.rm = TRUE))
    sldisc_rmse <- sqrt(mean((y_true - sldiscy_pred)^2, na.rm = TRUE))

    # Adding on RMSE from the single-best and ensemble
    rmse_df <- tibble(Learner = names(mean_rmse), RMSE = as.numeric(mean_rmse))
    superlearner_names <- c("Super Learner", "Discrete Learner")
    superlearner_rmse <- c(sl_rmse, sldisc_rmse)
    rmse_df <- bind_rows(rmse_df, tibble(Learner = superlearner_names, RMSE = superlearner_rmse))

    # Plotting RMSE performance
    slcv_plot <- ggplot(rmse_df, aes(x = reorder(Learner, RMSE), y = RMSE)) +
      scale_x_discrete(labels = pretty_labels) +
      geom_col(fill = "steelblue") +
      coord_flip() +
      theme_classic() +
      labs(title = paste("RMSE by Learner: ", target_var), x = "Learner", y = "RMSE")

    # Running normal SL for prediction with the same specifications
    sl_time <- system.time({
      slfull <- snowSuperLearner(
        Y = sldata[[target_var]],
        X = sldata[, toppreds],
        family = gaussian(),
        cluster = sl_cluster,
        SL.library = test.library,
        method = "method.NNLS",
        verbose = TRUE
      )
    })

    # Predicting with the SL ensemble, then rescaling predictions
    miss_data <- df_imp %>% filter(is.na(.data[[target_var]])) %>% select(all_of(toppreds))
    preds_scaled <- predict(slfull, newdata = miss_data, onlySL = TRUE)$pred[, 1]
    preds <- preds_scaled * range_y + min_y

    # Getting the residuals
    residuals_scaled <- slcv$SL.predict - slcv$Y
    
    # Setting resample count then resampling across all predicted values. Using simulate_resample function that uses nearby neighbors to help manage heteroskedasticity
    simulate_resample <- function(i, target_vals, residuals, preds, B = 1000) {
      local_resids <- residuals[order(abs(target_vals - preds[i]))[1:50]]
      preds[i] + sample(local_resids, size = B, replace = TRUE)
    }
    
    B <- 1000
    sim_mat <- future_sapply(seq_along(preds_scaled), simulate_resample, 
                             target_vals = slcv$Y, residuals = residuals_scaled, 
                             preds = preds_scaled, B = 1000, future.seed = TRUE)

    # Getting confidence intervals from that range, making sure ot scale back
    lower_ci <- apply(sim_mat, 2, quantile, probs = 0.025) * range_y + min_y
    upper_ci <- apply(sim_mat, 2, quantile, probs = 0.975) * range_y + min_y
    
    # Point median absolute deviation for weights, normalizing by the absolute mean of the resamples to help with the 0 to 1 scaling
    point_mads <- apply(sim_mat, 2, mad, constant = 1)
    point_means <- apply(sim_mat, 2, mean)
    rel_mads <- point_mads / (abs(point_means) + 1e-6)
    weights <- 1 / (1 + rel_mads)

    # Preparing data and then plotting for predicted distribution of resampled and exact predicted probabilities
    resampled_long <- data.frame(type = "Resampled", value = as.vector(sim_mat) * range_y + min_y)
    preds_long <- data.frame(type = "Predicted", value = preds)
    combined_data <- rbind(resampled_long, preds_long)

    pred_dist_plot <- ggplot(combined_data, aes(x = value, color = type, fill = type)) +
      geom_density(alpha = 0.3, size = 1) +
      theme_classic() +
      labs(title = paste0("Predicted vs Resampled Distributions: ", 
                          target_var), x = "Value", y = "Density", 
           color = "Distribution Type", fill = "Distribution Type")

    # Plotting ensemble coefficients (weights)
    slcv_coefs <- colMeans(slcv$coef)
    weights_df <- tibble(Learner = names(slcv_coefs), Weight = as.numeric(slcv_coefs))
    weight_plot <- ggplot(weights_df, aes(x = reorder(Learner, Weight), y = Weight)) +
      scale_x_discrete(labels = pretty_labels) +
      geom_col(fill = "steelblue") +
      coord_flip() +
      theme_classic() +
      labs(title = paste("SL Ensemble Weights:", target_var), x = "Learner", y = "Weights")

    # Preparing and then merging in the newly imputed data
    imp_final <- miss_data %>%
      mutate(.rowid = df_input %>% filter(is.na(.data[[target_var]])) %>% 
               pull(.rowid),
             !!paste0(target_var, "imp") := as.numeric(preds),
             !!paste0(target_var, "weight") := as.numeric(weights),
             !!paste0(target_var, "CI_lower") := as.numeric(lower_ci),
             !!paste0(target_var, "CI_upper") := as.numeric(upper_ci)) %>%
      select(.rowid, !!paste0(target_var, "imp"), ends_with("weight"), 
             ends_with("CI_lower"), ends_with("CI_upper"))

    obs_final <- df_input %>%
      filter(!is.na(.data[[target_var]])) %>%
      transmute(.rowid, !!paste0(target_var, "imp") := .data[[target_var]], 
                !!paste0(target_var, "weight") := 1, 
                !!paste0(target_var, "CI_lower") := .data[[target_var]], 
                !!paste0(target_var, "CI_upper") := .data[[target_var]])

    full_column <- bind_rows(obs_final, imp_final)

    df_augmented <- df_input %>%
      left_join(full_column, by = ".rowid") %>%
      select(-target_var)

    # Getting just the target var's results
    df_merged <- df_input %>%                     
      dplyr::left_join(full_column, by = ".rowid")
    
    imp_cols <- grep(paste0("^", target_var, "(imp|simimp)$"), names(df_merged), value = TRUE)
    aux_cols <- grep("(weight|CI_lower|CI_upper|lower_imp|upper_imp)$", names(df_merged), value = TRUE)
    
    keep_cols <- c(".rowid",
                   if (!is.null(id_var)) id_var,
                   target_var,
                   imp_cols,
                   aux_cols)
    keep_cols <- unique(keep_cols[keep_cols %in% names(df_merged)])
    
    var_data <- df_merged %>% dplyr::select(dplyr::all_of(keep_cols))

    # Returning list of diagnostics and data for user
    return(list(
      top10_plot = top10_plot, 
      mice_rmse = mice_rmse, 
      slcv_plot = slcv_plot, 
      sl_rmse = sl_rmse, 
      sldisc_rmse = sldisc_rmse, 
      sl_weights_plot = weight_plot, 
      prediction_distribution = pred_dist_plot, 
      updated_data = df_augmented, 
      var_data = var_data,
      high_missing = high_missing_flag, 
      mice_time_sec = mice_time["elapsed"], 
      sl_time_sec = sl_time["elapsed"]))
  }
}

### Multiple variable imputation
## Specifying the function
rs_multimvifunc <- function(target_vars, df_input, learners, screeners,
                            id_var = NULL, n_top_predictors = 10) {
  results <- list()
  current_df <- df_input
  n <- length(target_vars)

  ## Progress bar for long processes
  pb <- txtProgressBar(min = 0, max = n, style = 3)

  for (i in seq_along(target_vars)) {
    var <- target_vars[i]
    message(paste0("Processing ", var, " (", i, "/", n, ")..."))

    ## Distinguishing between binary and continuous input variables
    var_data <- current_df[[var]]
    var_type <- if (is.numeric(var_data) && length(unique(na.omit(var_data))) == 2) {
      "binary"
    } else {
      "continuous"
    }

    ## Suppressing messages/warnings and more (most/all are inconsequential and just flood the user)
    result <- withCallingHandlers(
      suppressWarnings(
        suppressMessages(
          {
            capture.output({
              result <- rs_mvifunc(
                target_var = var,
                df_input = current_df,
                learners = learners,
                screeners = screeners,
                var_type = var_type,
                id_var = id_var,
                n_top_predictors = n_top_predictors
              )
            }, type = "output")
            result
          }
        )
      ),
      warning = function(w) {
        if (!grepl("has >80% missingness", conditionMessage(w))) invokeRestart("muffleWarning")
      }
    )

    results[[var]] <- result
    current_df <- result$updated_data
    setTxtProgressBar(pb, i)
  }

  close(pb)

  ## Returning final data and all results
  return(list(
    all_results = results,
    final_data = current_df
  ))
}

### Specifying a test/default library of learners
lesslearners_both <- c("SL.bayesglm", "SL.caret.rpart", "SL.cforest",
                     "SL.glm", "SL.glm.interaction",
                     "SL.glmnet", "SL.kernelKnn", "SL.ksvm", 
                     "SL.mean", "SL.nnet", "SL.randomForest", 
                     "SL.ranger", "SL.rpart", "SL.rpartPrune", "SL.speedglm",
                     "SL.step", "SL.step.forward", "SL.stepAIC",
                     "SL.xgboost")
```

### Generating synthetic dataset

The synthetic dataset consisted of 1000 subjects and 100 features and
was generated to support this illustration. It includes simulated
missingness relevant to real-world data. Feature generation included
predefined domains of continuous laboratory value variables (LAB),
binary Next-Generation Sequencing variables (NGS), binary mutation flag
variables (MUTATION), and binary cytometry variables (CYT) simply
numbered iteratively. 70% of variables were targeted to be binary and
made of the CYT, NGS, and MUTATION domains. Random inter-feature
correlations and missingness patterns were introduced resulting in a
range of target missingness contexts.

### Single binary variable imputation function

When applying the `rs_multimvifunc` there are many outputs
that allow a user to understand the performance of not only the SL-based
approach but also how it compares to a standard MICE approach.
Furthermore, there are chances to understand how the SL ensemble was
assembled. The primary output of the SL-based function is an updated
dataset that has imputed values for all missing values of all target
variables. Primary output also includes 0-1 scaled uncertainty weights
of those imputations (with observed data retaining a weight of 1) as a
quantification of how certain the imputations were. Binary variable
output of the SL-based imputation function includes:

`updated_data`: the updated dataset that has the newly
created imputed variables and weights merged into the input dataset. It
is important to note if the user wants summary statistics of the weights
for the imputations alone, the user will need to filter out all
observations with a weight of 1.

`top10_plot`: a plot of the top 10 predictors (or however
many predictors were specified) and their respective predictor scores
(calculated via a formula that balances predictor missingness with a
predictor’s correlation with a given target variable). With this plot
users can understand what variables contributed most to predicting a
given target variable.

`mice_confmatrix`: a confusion matrix and related
performance measures, including balanced accuracy, for the MICE-based
approach. The performance measures reported in this object give a
comprehensive understanding of MICE’s imputation performance.

`mice_f1`: the MICE approach’s F1 score.

`sl_confusion`: a confusion matrix and related performance
measures for the SL ensemble approach, with similar utility to MICE’s
confusion matrix and related performance measures.

`sldisc_confusion`: a confusion matrix and related
performance measures for the single best-performing learner from the SL
library.

`sl_resample_ba`: the resampling-derived balanced accuracy
and 95% confidence intervals for the SL-based approach. This allows
users to have a measure of uncertainty regarding a key binary variable
imputation performance measure.

`sl_f1`: the SL-based approach’s F1 score.

`sl_table`: a 2x2 table class object for the SL-based
approach, enabling users to manually calculate other SL-based
performance measures.

`slcv_plot`: performance, measured as AUC for binary
variables, for all individual algorithms averaged across all
cross-validation loops of the SL process. This plot informs users of how
individual algorithms performed on the data, on average. The simple mean
learner will ideally perform the worst, otherwise something may be wrong
with the configuration or the data.

`sl_weights_plot`: a plot showing the average ensemble
weights assigned to individual algorithms across all iterations of the
SL cross-validation loop. As described in the manuscript, the SL
approach will find the most optimal combination of algorithms to yield
the highest performance, as measured by AUC for binary variables. This
will not always be the highest performers seen on the
`slcv_plot`.

`opt_threshold`: the optimal threshold selected by the
SL-based approach to classify predicted values by. This is selected via
AUC when class imbalance is less than 90%, or F1 score when class
imbalance is greater than or equal to 90% but lower than 95%

`prediction_distribution`: the predicted probability
density distribution from the prediction of true new missing data,
overlaid with the resampled predicted probability distribution to
indicate potential variability.

`high_missing`: a true/false flag indicating if the target
variable was missing greater than 80% of its data, in which case
imputations should be taken with high caution.

`mice_time_sec`: the computational run time of MICE in
seconds.

`sl_time_sec`: the computational run time of the SL
ensemble trained for prediction in seconds, which is most comparable to
MICE. This is not the computational time for the entire SL-based
function or the SL cross validation loop that generates performance
measures.

`var_data`: just the given target variable’s
imputation-relevant data (imputed variable with carried forward observed
values, uncertainty weight, lower and upper possible imputation
values).

Observing the aforementioned output for a given target variable,
CYT\_13, we can understand that the SL-based imputation function yields
higher performance. We can additionally investigate how the SL
approach’s individual algorithms performed on average, how the SL
approach assigned algorithm weights, what the optimal threshold for
classification was, and the distribution of the predicted and resampled
values.

```
# Printing outputs
rs_mvifunc_bintest$all_results$CYT_13$top10_plot
```

```
## MICE vs SL performance by balanced accuracy
get_ba <- function(cm) {
  if (!is.null(cm$byClass) && "Balanced Accuracy" %in% names(cm$byClass)) {
    as.numeric(cm$byClass["Balanced Accuracy"])
  } else if (!is.null(cm$table)) {
    tn <- cm$table[1,1]; fp <- cm$table[1,2]
    fn <- cm$table[2,1]; tp <- cm$table[2,2]
    sens <- tp/(tp+fn); spec <- tn/(tn+fp)
    mean(c(sens, spec), na.rm = TRUE)
  } else NA_real_
}

# Extract values
mice_ba  <- get_ba(rs_mvifunc_bintest$all_results$CYT_13$mice_confmatrix)
sl_ba_cv <- get_ba(rs_mvifunc_bintest$all_results$CYT_13$sl_confusion)

ba_vec <- as.numeric(rs_mvifunc_bintest$all_results$CYT_13$sl_resample_ba)
sl_ba_boot <- ba_vec[1]
sl_ba_lcl  <- ba_vec[2]
sl_ba_ucl  <- ba_vec[3]

# Build summary table
tibble(
  Method = c("MICE", "SL (CV)", "SL (resampled)"),
  `Balanced Accuracy` = c(mice_ba, sl_ba_cv, sl_ba_boot),
  `95% CI` = c(NA, NA, sprintf("[%.3f, %.3f]", sl_ba_lcl, sl_ba_ucl))
) %>%
  kable(digits = 3, caption = "Balanced Accuracy (BA) Summary")
```

Balanced Accuracy (BA) Summary

| Method | Balanced Accuracy | 95% CI |
| --- | --- | --- |
| MICE | 0.521 | NA |
| SL (CV) | 0.694 | NA |
| SL (resampled) | 0.693 | [0.658, 0.725] |

```
rs_mvifunc_bintest$all_results$CYT_13$slcv_plot
```

```
rs_mvifunc_bintest$all_results$CYT_13$sl_weights_plot
```

```
data.frame(`Optimal Threshold` = round(rs_mvifunc_bintest$all_results$CYT_13$opt_threshold, 3)) %>%
  kable(caption = "Optimal classification threshold (SL-based)")
```

Optimal classification threshold (SL-based)

Optimal.Threshold

0.27

```
rs_mvifunc_bintest$all_results$CYT_13$prediction_distribution
```

```
data.frame(
  Method = c("MICE time (sec)", "Super Learner time (sec)"),
  Time_sec = c(
    rs_mvifunc_bintest$all_results$CYT_13$mice_time_sec, 
    rs_mvifunc_bintest$all_results$CYT_13$sl_time_sec
  )
) %>%
  kable(caption = "Computation Times for CYT_13")
```

Computation Times for CYT\_13

| Method | Time\_sec |
| --- | --- |
| MICE time (sec) | 0.737 |
| Super Learner time (sec) | 88.270 |

### Single continuous variable imputation function

Applying `rs_multimvifunc` for a continuous target
variable outputs similar objects that enable users to evaluate SL-based
performance, compare it to a MICE benchmark, and explore how the SL
ensemble was constructed. The updated dataset returned includes imputed
values for the target variable and 0–1 scaled uncertainty weights, with
observed data assigned a weight of 1. For these continuous variables,
outputs additionally include 95% observation-specific confidence
intervals for different lower and upper assumptions in downstream
analyses of the imputed values. Root Mean Square Error (RMSE) measures
performance, which can be interpreted in that value’s original units.
Continuous variable output of the SL-based imputation function
includes:

`updated_data`: the updated dataset containing the imputed
continuous variable, its uncertainty weights, and associated confidence
intervals merged into the input dataset. Users should filter out weights
equal to 1 to summarize uncertainty among imputations only.

`top10_plot`: visualizes the top 10 predictors and their
predictor scores, reflecting a balance between correlation with the
target variable and completeness.

`mice_rmse`: RMSE of the MICE approach, summarizing
imputation performance for the continuous variable.

`sl_rmse`: RMSE of the full SL ensemble.

`sldisc_rmse`: RMSE for the single best-performing learner
from the SL library.

`sl_weights_plot`: displays the ensemble weights assigned
to each learner by the SL process, which may differ from the learners’
individual rankings in slcv\_plot.

`prediction_distribution`: shows the distribution of
predicted continuous values alongside their resampled distributions from
the bootstrap procedure, reflecting imputation variability.

`high_missing`: a flag indicating if the target variable
had greater than 80% missingness.

`mice_time_sec`: computational runtime of the MICE
procedure.

`sl_time_sec`: computational runtime for the SL ensemble
trained for prediction, most directly comparable to
`mice_time_sec`.

`var_data`: just the given target variable’s
imputation-relevant data (imputed variable with carried forward observed
values, uncertainty weight, lower and upper possible imputation
values).

For a given continuous target variable, reviewing these outputs
allows users to again determine which predictors were most influential,
compare ensemble versus individual learner performance, evaluate
imputation uncertainty via both weights and confidence intervals, and
assess runtime.

```
# Printing outputs
rs_mvifunc_conttest$all_results$LAB_06$top10_plot
```

```
data.frame(
  Method = c("MICE", "SL-based"),
  RMSE = c(
    rs_mvifunc_conttest$all_results$LAB_06$mice_rmse,
    rs_mvifunc_conttest$all_results$LAB_06$sl_rmse
  )
) %>%
  kable(caption = "RMSE Comparison for LAB_06")
```

RMSE Comparison for LAB\_06

| Method | RMSE |
| --- | --- |
| MICE | 8.829954 |
| SL-based | 6.375208 |

```
rs_mvifunc_conttest$all_results$LAB_06$slcv_plot
```

```
rs_mvifunc_conttest$all_results$LAB_06$sl_weights_plot
```

```
rs_mvifunc_conttest$all_results$LAB_06$prediction_distribution
```

```
data.frame(
  Method = c("MICE time (sec)", "Super Learner time (sec)"),
  Time_sec = c(
    rs_mvifunc_conttest$all_results$LAB_06$mice_time_sec, 
    rs_mvifunc_conttest$all_results$LAB_06$sl_time_sec
  )
) %>%
  kable(caption = "Computation Times for LAB_06")
```

Computation Times for LAB\_06

| Method | Time\_sec |
| --- | --- |
| MICE time (sec) | 0.491 |
| Super Learner time (sec) | 56.493 |

### Vector of variables imputation function

This section demonstrates applying the multi-variable SL-based
imputation function to a vector of variables, automatically ordering
them from least to most missing before sequential imputation. The output
provides updated data with imputations and uncertainty weights for each
variable again, along with key performance measures for both the
SL-based and MICE approaches. This allows side-by-side comparison of
performance across multiple variables in a single streamlined run. For
brevity, we briefly summarize the key performance measures across this
application to three binary variables in the synthetic data.

```
# Looking at just a few performance measures
metrics_summary <- map_dfr(names(rs_multimvifunctest$all_results), function(var) {
  res <- rs_multimvifunctest$all_results[[var]]
  tibble(
    Variable              = var,
    MICE_Balanced_Accuracy = 100 * res$mice_confmatrix$byClass["Balanced Accuracy"],
    SL_Balanced_Accuracy   = 100 * res$sl_confusion$byClass["Balanced Accuracy"],
    MICE_F1                = 100 * res$mice_f1,
    SL_F1                  = 100 * res$sl_f1,
    MICE_Time_Sec          = res$mice_time["elapsed"],
    SL_Time_Sec            = res$sl_time_sec
  )
})

metrics_sumsum <- metrics_summary %>%
  summarise(
    `MICE (%)`   = mean(MICE_Balanced_Accuracy, na.rm = TRUE),
    `SL (%)`     = mean(SL_Balanced_Accuracy,   na.rm = TRUE),
    `MICE F1 (%)`= mean(MICE_F1,                na.rm = TRUE),
    `SL F1 (%)`  = mean(SL_F1,                  na.rm = TRUE),
    `MICE (sec)` = mean(MICE_Time_Sec,          na.rm = TRUE),
    `SL (sec)`   = mean(SL_Time_Sec,            na.rm = TRUE)
  ) %>%
  mutate(across(everything(), ~ round(., 1))) %>%
  mutate(
    `MICE (%)`    = sprintf("%.1f%%", `MICE (%)`),
    `SL (%)`      = sprintf("%.1f%%", `SL (%)`),
    `MICE F1 (%)` = sprintf("%.1f%%", `MICE F1 (%)`),
    `SL F1 (%)`   = sprintf("%.1f%%", `SL F1 (%)`)
  )

df <- as.data.frame(metrics_sumsum)
rownames(df) <- rep("", nrow(df))

kbl(df,
    caption = "Summary of Performance Across Variables",
    align = "c",
    booktabs = TRUE,
    row.names = TRUE) |>
  add_header_above(c(" " = 1, "Balanced Accuracy" = 2, "F1" = 2, "Runtime" = 2)) |>
  kable_styling(full_width = FALSE,
                bootstrap_options = c("striped","condensed","hover"),
                position = "left")
```

Summary of Performance Across Variables

|  | Balanced Accuracy | | F1 | | Runtime | |
| --- | --- | --- | --- | --- | --- | --- |
|  | MICE (%) | SL (%) | MICE F1 (%) | SL F1 (%) | MICE (sec) | SL (sec) |
|  | 59.7% | 70.0% | 73.3% | 80.0% | 0.4 | 42.7 |
